# Leveraging Generative Artificial Intelligence for Enhanced Data Augmentation in Emotion Intensity Classification: A Comprehensive Framework for Cross-Dataset Transfer Learning

**DOI:** 10.64898/2026.02.23.26346928

**Authors:** Julia Wieczorek, Xiaorui Jiang, Vasile Palade, Joanna Trela

**Affiliations:** Coventry University, Priory Street, Coventry, CV1 5FB, United Kingdom; University of Sheffield, Department of Computer Science, Regent Court, 211 Portobello, Sheffield S1 4DP, United Kingdom

## Abstract

Data scarcity and stylistic heterogeneity pose major challenges for emotion intensity classification. This paper presents a cross-dataset augmentation framework that leverages prompt-conditioned generative models alongside deterministic and heuristic transformations to synthesize target-style examples for improved transfer learning. We introduce a unified taxonomy of augmentation strategies—Heuristic Lexical Perturbation (HLA), Prompt-Conditioned Generative Augmentation (CGA), Sequential Hybrid Pipeline (SHA), Rule-Guided Style Adaptation (DSGA), and Enhanced Hybrid Augmentation (EHA)—and detail an interpretability-oriented prompt engineering approach that conditions LLMs on authentic target exemplars and stylistic features extracted from the target dataset.

Augmented datasets were evaluated using multi-dimensional quality metrics (transformation quality, stylistic consistency, BLEU/CHRF, Self-BLEU, uniqueness) and downstream classification via a two-phase BERT-LSTM training with rigorous statistical testing. During source dataset pretraining and subsequent target dataset fine-tuning, CGA achieved the highest single-method gains in F1 and accuracy (F1 = 0.8816; accuracy = 0.8819, 95% CI recalculated). HLA and SHA exhibited improved cross-domain stability, suggesting stronger domain-generalizable features. We observe systematic trade-offs between fluency, lexical diversity, and emotion fidelity: high surface similarity often correlates with classifier performance but does not fully capture affective authenticity.

We discuss methodological pitfalls, propose best practices for emotion-aware augmentation, and provide reproducible artifacts (prompts, example transformations, evaluation scripts) to facilitate further research in affective NLP.

## 1 Introduction

Deep learning models work best when they have access to large, high-quality datasets. In some natural language processing tasks, this creates a significant challenge because obtaining enough labeled data for supervised learning tasks is often difficult and expensive. The shortage of annotated datasets directly limits how well models can perform, while creating large-scale datasets manually requires considerable time and resources [Silva et al., 2023].

However, augmenting text data is more complex than augmenting other types of data. Language has intricated grammatical rules, semantic meanings, and contextual relationships that make it hard to create synthetic text that maintains the original quality. Research shows that poorly designed text augmentation can actually hurt model performance, especially when the generated text contains grammatical errors or changes the intended emotional meaning [Luo et al., 2021]. Simply copying existing data does not solve the problem and can lead to overfitting, making it clear that better approaches to synthetic text generation are needed.

These issues are particularly important for emotion intensity classification, where maintaining the authenticity of emotional expression is crucial. This task plays a key role in intelligent systems used for human-computer interaction, mental health support, and social media analysis. Being able to accurately measure how intense emotions are in text helps create AI systems that can respond more appropriately to human feelings [Akhtar et al., 2020]. Traditional augmentation methods often fail to preserve the subtle emotional characteristics that make human expression authentic, which can reduce the effectiveness of trained models [Zhang and Xu, 2021].

Recent developments in large language models provide new opportunities to address these challenges. They can understand context and generate coherent text that preserves both style and emotional authenticity [Brown et al., 2020, Touvron et al., 2023a]. This capability is especially valuable for emotion intensity classification, where keeping the original emotional meaning intact is essential for training effective models.

This chapter examines how generative AI can be integrated into data augmentation frameworks specifically for emotion intensity classification in intelligent systems. We focus on answering this key question: How can large language models be used to create high-quality synthetic training data that maintains both linguistic authenticity and emotional coherence?

Our research combines text generation capabilities of LLMs with careful quality control measures to create augmented datasets that improve classification performance without the typical drawbacks of synthetic data. The results provide insights into using generative AI in intelligent systems and offer a practical framework for applying LLM-based data augmentation to other natural language processing tasks where limited data presents significant challenges.

## 2 Background and Related Work

### 2.1 Data Augmentation in Natural Language Processing

Data augmentation (DA) is a strategy to increase a system’s performance by generating more training data. It can significantly enhance the effectiveness of cross-dataset transfer learning by addressing the specific challenges that arise when adapting models between different emotion intensity datasets. Rather than relying solely on direct transfer of learned representations, augmentation-supported transfer creates synthetic bridging data that helps models adapt to new domains while preserving valuable knowledge from source datasets.

DA encompasses methods of increasing training data diversity without directly collecting more data, with the goal of having the augmented data act as a regularizer to reduce overfitting when training machine learning models [Shorten and Khoshgoftaar, 2019]. While data augmentation has proven highly effective in computer vision through techniques such as image rotation, cropping, and color adjustments, natural language processing presents unique challenges due to the discrete and structured nature of language.

The theoretical understanding of why data augmentation works remains limited. As noted by Dao et al. [2019], data augmentation is typically performed in an ad-hoc manner with little understanding of the underlying theoretical principles, and the typical explanation of data augmentation as regularization may be insufficient. Some research suggests that training with noised examples can be reduced to Tikhonov regularization [Bishop, 1995], while other studies highlight benefits such as feature averaging and variance reduction [Cheng et al., 2020]. Despite these theoretical insights, the fundamental mechanisms of augmentation in NLP remain underdeveloped, particularly in specialized domains where semantic preservation is critical.

#### 2.1.1 Traditional Augmentation Approaches

Traditional NLP augmentation methods can be categorized into three main approaches: rule-based techniques (Easy Data Augmentation (EDA)), example interpolation methods (MIXUP [Zhang et al., 2017], SEQ2MIXUP [Guo, 2020]), and model-based approaches (Seq2seq, language model, fine-tuning GPT-2, paraphrasing). Rule-based methods include synonym replacement, random word insertion and deletion, sentence shuffling, and back-translation [Wei and Zou, 2019, Feng et al., 2021]. These techniques are easy to implement but often provide only modest performance improvements. Moreover, simple lexical substitutions can inadvertently alter semantic meaning, particularly in specialized domains where subtle linguistic cues carry significant weight [Feng et al., 2021].

Example interpolation methods, such as MIXUP adaptations for text, blend features from multiple examples to create synthetic training data [Zhang et al., 2017]. However, these approaches face inherent limitations in discrete language spaces where interpolation between word embeddings may not correspond to meaningful linguistic expressions. Early model-based approaches leveraged trained language models to generate new examples, offering more sophisticated augmentation but requiring additional computational resources.

The effectiveness of text augmentation depends heavily on maintaining the balance between increasing data diversity and preserving semantic integrity. The discrete nature of language makes it difficult to apply continuous transformations that work well in other domains, and simple modifications can inadvertently change the intended meaning of text.

#### 2.1.2 Challenges in Emotion-Related Tasks

This challenge is particularly difficult in emotion-related tasks, where subtle linguistic features and contextual relationships are crucial for conveying emotional intensity. Word-level changes that seem minor may significantly alter the emotional content, potentially creating mislabeled synthetic samples that harm rather than help model performance [Mohammad and Bravo-Marquez, 2017].

Emotion intensity datasets typically exhibit severe imbalances across intensity levels, with extreme intensities (both very low and very high) being significantly underrepresented compared to moderate levels. Rule-based techniques such as EDA – including synonym replacement, random insertion, deletion, and sentence shuffling – offer computational efficiency and simplicity but present significant risks for emotion intensity tasks. EDA’s random lexical substitutions can inadvertently alter emotional intensity in unpredictable ways: replacing “devastated” with “sad” maintains emotional valence but substantially reduces intensity, while substituting “quite upset” with “extremely upset” through random insertion can artificially inflate intensity levels. These uncontrolled transformations often generate training samples with misaligned intensity labels, potentially degrading model performance rather than improving it.

The discrete nature of emotional language compounds these challenges. Unlike computer vision where geometric transformations preserve object identity, linguistic transformations in EDA operate without semantic awareness. Random word deletion may remove crucial intensity modifiers (“very,” “extremely,” “slightly”), while random insertion may introduce contradictory emotional signals. Back-translation, while more semantically aware than pure lexical methods, still struggles to preserve fine-grained intensity distinctions across language boundaries.

Model-based augmentation techniques address many limitations of traditional approaches by maintaining semantic and emotional coherence during transformation. These models can generate emotionally consistent variations that preserve both the intended emotional content and its intensity level while introducing beneficial linguistic diversity. However, LLM-based approaches require careful prompt engineering and quality control to ensure generated samples maintain appropriate intensity-label alignment.

### 2.2 Pre-trained Language Models and Generation Capabilities

Generative Artificial Intelligence in natural language processing refers to AI systems capable of creating text, understanding context, and generating human-like responses through sophisticated generative models that learn patterns and structures from training data [Sengar et al., 2025]. The development of large-scale pre-trained language models has fundamentally transformed natural language processing capabilities, particularly in the context of text generation and augmentation.

#### 2.2.1 Evolution from Static to Contextual Representations

The transformer architecture, introduced by Vaswani et al. [2017], laid the foundation for subsequent developments in generative AI, enabling models to capture long-range dependencies and contextual relationships in text with remarkable accuracy. Early approaches to text representation, such as word2vec [Mikolov et al., 2013] and GloVe [Pennington et al., 2014], provided static embeddings that captured semantic relationships but failed to account for contextual variation.

The introduction of ELMo [Peters et al., 2018] marked a shift toward contextual representations, where word meanings adapt based on surrounding context. BERT (Bidirectional Encoder Representations from Transformers) [Devlin et al., 2019] further advanced contextual understanding through bidirectional processing, enabling models to capture complex semantic relationships for downstream tasks. BERT represents a significant advancement in pre-training techniques for NLP tasks, designed to capture contextual information bidirectionally from both left and right sides of words in sentences. After pre-training, BERT can be fine-tuned for specific downstream tasks such as sentiment analysis, question answering, and named entity recognition.

#### 2.2.2 Large Language Models and Controlled Generation

GPT models are built on transformer architectures specifically designed for generating coherent and contextually relevant text [Radford et al., 2019]. These models demonstrate exceptional capabilities in text generation tasks, leveraging pre-training on massive datasets to understand language patterns and generate human-like responses across diverse contexts. More recent models such as LLaMA [Touvron et al., 2023a] have further demonstrated that scale and architectural improvements enable more sophisticated text generation capabilities.

The emergence of instruction-following capabilities in large language models has opened new possibilities for controlled text generation in data augmentation contexts. Unlike traditional rule-based methods, these models can understand high-level instructions about desired transformations while maintaining semantic coherence and stylistic authenticity, making them particularly suitable for emotion-related applications where preserving affective nuance is essential.

#### 2.2.3 Applications to Data Augmentation

Modern language models enable several augmentation paradigms not feasible with traditional approaches. Prompt-based generation allows researchers to specify desired characteristics of synthetic data, such as emotional tone, stylistic patterns, or domain-specific vocabulary. Cross-domain adaptation represents another significant capability, where models can transform text from one domain to match the stylistic and semantic characteristics of another domain, particularly valuable for addressing data scarcity in specialized domains by leveraging knowledge from related but more abundant datasets.

However, these capabilities also introduce new challenges. Ensuring consistency between generated text and assigned labels becomes critical, as sophisticated generation capabilities may produce text that appears plausible but misaligns with intended semantic categories [Ding et al., 2024].

### 2.3 Emotion Intensity Classification

Emotion intensity classification is a subfield of affective computing that aims to quantify the strength or degree of emotional expression in textual data. Unlike traditional sentiment analysis, which focuses primarily on the polarity of emotions, emotion intensity classification seeks to determine how strongly an emotion is expressed [Oladepo et al., 2025].

#### 2.3.1 Task Definition

Emotion intensity classification involves determining how strongly an emotion is expressed in text, typically on ordinal or continuous scales. This task proves challenging due to the subjective nature of emotional expression and the cultural, contextual, and individual factors that influence emotional communication patterns.

The limited availability of high-quality emotion intensity datasets compounds these challenges. Annotation requires consistent interpretation of emotional expression across annotators, and many existing datasets suffer from class imbalance, particularly underrepresentation of extreme intensity levels. Crossdomain generalization adds another layer of difficulty, as emotional expression patterns vary significantly across contexts, platforms, and communities.

#### 2.3.2 Annotation Inconsistences and Labeling Challenges

A fundamental challenge in emotion intensity classification stems from the lack of standardized annotation approaches across datasets. This inconsistency manifests in multiple dimensions: scaling systems, annotation methodologies, and the philosophical foundations underlying emotional labeling.

Different datasets employ varying intensity scales that complicate cross-dataset comparison and model transfer. For instance, MEISD [Firdaus et al., 2020] utilizes a 0-3 discrete scale for intensity ratings, while ESConv [Liu et al., 2021] employs a discrete 1-5 scale. This scale heterogeneity requires careful normalization procedures when combining datasets or transferring models between domains, as the same numerical intensity value may represent different emotional magnitudes across datasets.

The annotation methodologies vary substantially between datasets, reflecting different theoretical approaches to emotional assessment. With the same dataset example, annotation in MEISD was performed by highly proficient in English comprehension graduate students who utilized transcripts, video clips, and audio recordings to ensure precise labeling, thereby addressing the inherent ambiguity in text-only emotion recognition. In contrast, ESConv annotations were provided by the same individuals who experienced the emotions and participated in the conversations, resulting in more subjective but deeply grounded emotional labels that reflect the speaker’s internal state. This difference highlights a key distinction in annotation philosophy between the two datasets—objective external observation versus self-reported emotional experience. While external annotation may provide more consistent inter-rater reliability, self-reported annotations capture the authentic emotional experience of the speaker, albeit with potential subjectivity biases.

#### 2.3.3 Domain Adaptation Challenges

Emotion intensity models frequently encounter domain shift when deployed across different contexts or datasets. Expression patterns that indicate high emotional intensity in one domain may represent moderate intensity in another, and vocabulary choices, sentence structures, and cultural references can vary significantly between domains.

Texts originating from different sources exhibit distinct characteristics that complicate intensity classification. Social media posts typically feature informal language, abbreviations, and emotional amplification through repetitive punctuation or capitalization. Conversational dialogues maintain interpersonal dynamics and contextual dependencies that influence emotional expression. Content from TV series reflects scripted emotional performances that may exaggerate or stylize natural emotional expression. Therapeutic conversations with psychologists involve professional communication norms that may constrain emotional expression compared to peer-to-peer interactions.

Different digital platforms foster unique communication cultures that affect emotional expression intensity. Chat-based interactions with automated systems may elicit more direct emotional expression, while conversations with human therapists involve social desirability biases that moderate emotional intensity reporting. Social media platforms encourage performative emotional expression, while private therapeutic settings promote authentic but potentially guarded emotional disclosure.

The ESConv dataset [Liu et al., 2021] and MEISD dataset [Firdaus et al., 2020] exemplify these domain differences—emotional support conversations versus TV series dialogues—requiring models to adapt to distinct communicative norms, intensity scales, and stylistic conventions. This domain adaptation challenge motivates the need for augmentation techniques that can bridge these gaps while preserving emotional authenticity.

Emotional expression patterns vary significantly across cultural and demographic groups, adding complexity to cross-domain adaptation. Collectivist cultures may exhibit more moderate individual emotional expression compared to individualist cultures, while age and gender demographics influence both vocabulary choices and intensity expression patterns. These cultural variations necessitate careful consideration when developing emotion intensity models for diverse populations or when transferring models across different demographic contexts.

#### 2.3.4 Applications

##### Mental Health and Crisis Intervention

Emotion intensity classification plays a critical role in automated mental health monitoring systems. For instance, the detection of high-intensity expressions of despair, anxiety, or hopelessness in social media posts or therapy chat logs can trigger early intervention mechanisms. Studies have shown that individuals experiencing suicidal ideation often exhibit escalating emotional intensity in their written communications days or weeks before crisis events [Coppersmith, 2018]. Automated systems capable of detecting these intensity patterns have been deployed in platforms like Crisis Text Line, where they help prioritize urgent cases among thousands of daily conversations.

##### Therapeutic Support Systems

In digital therapy platforms such as Woebot or Wysa, emotion intensity classification enables personalized response generation. When a user expresses moderate anxiety, the system might suggest breathing exercises, but high-intensity panic expressions trigger more immediate coping strategies or human therapist referrals. The ESConv dataset [Liu et al., 2021] specifically captures these nuanced therapeutic interactions, where counselors adjust their intervention strategies based on the perceived emotional intensity of help-seekers’ messages.

##### Customer Experience and Brand Management

Beyond clinical applications, emotion intensity classification has significant commercial applications. Companies use these systems to prioritize customer service responses, with high-intensity anger or frustration receiving immediate escalation to human agents. Social media monitoring tools employ intensity classification to distinguish between mild dissatisfaction (manageable through automated responses) and viral complaint potential (requiring immediate damage control).

##### Educational Technology

Adaptive learning systems increasingly incorporate emotion intensity analysis to optimize student engagement. High-intensity frustration signals may trigger simplified explanations or additional practice problems, while expressions of boredom might prompt more challenging or gamified content. Research in educational psychology demonstrates that learning outcomes improve significantly when instruction adapts to emotional state intensity rather than merely content comprehension levels [Liu et al., 2024, Wu and Yu, 2022].

### 2.4 Sequential Transfer Learning

Sequential transfer learning addresses the fundamental challenge of limited training data in emotion intensity classification by leveraging knowledge acquired from related tasks or domains. This paradigm proves particularly valuable in affective computing, where high-quality annotated datasets are scarce and expensive to produce. Rather than training models from scratch for each new dataset or domain, sequential transfer learning enables models to build upon previously learned representations of emotional expression patterns.

However, the challenges identified in emotion intensity classification—annotation inconsistencies, scale variability, and domain adaptation difficulties—create unique complications for transfer learning approaches. The heterogeneity in labeling methodologies and intensity scales means that knowledge transfer between datasets requires careful calibration to account for systematic differences in how emotional intensity is measured and interpreted across different annotation frameworks.

#### 2.4.1 Cross-Dataset Transfer

Cross-dataset transfer in emotion intensity classification involves training models on source datasets and adapting them to target datasets with potentially different annotation schemes, intensity scales, and domain characteristics. This approach addresses the practical reality that most emotion intensity datasets are small-scale and domain-specific, making it necessary to leverage knowledge from multiple sources to achieve robust performance.

##### Scale Alignment Challenges

The variability in intensity scales across datasets presents immediate technical challenges for cross-dataset transfer. When transferring from continuous scale to discrete scale, models must learn to map between fundamentally different numerical representations of emotional intensity. Simple linear transformations often prove insufficient, as the underlying intensity distributions may follow different patterns across datasets.

##### Annotation Philosophy Mismatch

The philosophical differences in annotation approaches create additional complexity for cross-dataset transfer. Models trained on externally-annotated data learn to recognize emotional intensity as perceived by objective observers, while models trained on self-reported annotations learn to predict the speaker’s internal emotional state. These represent fundamentally different learning objectives that may not transfer seamlessly. External annotations may focus on linguistic markers and contextual cues visible to third parties, while self-reported annotations capture subtle internal experiences that may not manifest clearly in textual expression.

##### Domain-Specific Features

Cross-dataset transfer must account for domain-specific vocabulary, communication patterns, and contextual factors that influence emotional expression. Models trained on social media data may overweight certain linguistic features (such as capitalization or repetitive punctuation) that are less relevant in therapeutic conversation contexts. Conversely, models trained on formal therapeutic dialogues may underperform on informal social media expressions where emotional intensity is conveyed through different linguistic mechanisms.

##### Evaluation Challenges

Assessing the success of cross-dataset transfer requires careful consideration of evaluation metrics that account for scale differences and annotation philosophies. Standard metrics may not adequately capture transfer performance when intensity scales and distributions differ significantly between source and target datasets. Some research employs rank-based metrics or correlation measures that are more robust to scale transformations, while others develop dataset-specific evaluation frameworks that account for annotation methodology differences.

#### 2.4.2 Augmentation-Supported Transfer Learning

Augmentation-supported transfer learning represents a synergistic approach that combines the knowledge preservation capabilities of transfer learning with the data diversity and domain adaptation benefits of data augmentation. Rather than viewing these techniques as separate methodologies, this paradigm integrates them into a unified framework that addresses the multifaceted challenges of cross-dataset emotion intensity classification.

In augmentation-supported transfer, during the pre-transfer phase, augmentation can enhance source dataset coverage and balance, ensuring that transferred knowledge represents a comprehensive understanding of emotional intensity patterns. During the transfer phase, augmentation can generate bridging samples that facilitate smoother domain adaptation by creating intermediate representations between source and target domains. Finally, during the post-transfer phase, targeted training can address specific gaps or biases in the target domain while preserving valuable knowledge acquired from the source domain.

This process can generate synthetic samples that interpolate between source and target domain characteristics, creating smoother transitions for transfer learning. For example, when transferring from social media to therapeutic conversation domains, this process gradually shifts from informal social media language patterns to more formal therapeutic communication styles. This bridging approach helps models adapt incrementally rather than requiring abrupt domain shifts that may lead to performance degradation.

The integration of data augmentation with transfer learning represents a promising approach for addressing the multi-faceted challenges of emotion intensity classification. By combining the knowledge preservation capabilities of transfer learning with the data diversity and domain adaptation benefits of augmentation, this hybrid approach can achieve more robust and generalizable emotion intensity models that perform effectively across diverse datasets and application contexts.

### 2.5 Quality Assessment in Synthetic Data Generation

As synthetic data generation becomes increasingly integrated into natural language processing (NLP) workflows, particularly through large language models (LLMs), the need for systematic quality evaluation becomes critical. Traditional evaluations often rely on model performance metrics such as accuracy or F1-score after training on augmented datasets. However, these downstream metrics alone are insufficient to assess the intrinsic quality, diversity, and utility of the generated text.

#### 2.5.1 Multi-Dimensional Quality Assessment

To address this gap, recent work by Amadeus and Cruz Castañeda [2024] proposes a taxonomy of evaluation metrics for text data augmentation, aiming to provide a unified benchmark for both academic and industrial contexts. Their taxonomy categorizes quality assessment into several key dimensions:

Fluency and Semantic Coherence metrics such as Perplexity (PPL) and syntactic log-odds ratio (SLOR) [Kann et al., 2018] evaluate how linguistically natural the generated samples are, while semantic fidelity is often measured through embedding-based similarity or content preservation scores. Diversity and novelty metrics such as Self-BLEU, Unique Trigram Ratio (UTR), and Type-Token Ratio (TTR) capture how distinct the synthetic samples are, both from each other and from the original dataset, thus mitigating risks of redundancy or overfitting.

Style and Sentiment Consistency becomes particularly important for emotionally grounded tasks such as emotion intensity classification, where it is essential that augmented texts maintain emotional tone. Metrics like Sentiment Difference (SENTDIFF) and Sentiment Standard Deviation (SENTSTD) assess whether transformations preserve or shift emotional polarity.

Textual Similarity and Alignment can be measured through character-based metrics such as CHRF (Character n-gram F-score) and BLEU, with CHRF being particularly effective in morphologically rich or low-resource languages.

#### 2.5.2 Emotion-Specific Quality Considerations

For emotion-related tasks, maintaining affective authenticity is critical. Standard NLP evaluation metrics may indicate high lexical similarity while missing important shifts in emotional tone or intensity that could mislead classification models. This limitation necessitates the development of more sophisticated quality assessment frameworks that incorporate emotion-specific evaluations alongside traditional metrics.

Incorporating these dimensions into a comprehensive quality assessment framework enables not only a more nuanced evaluation of synthetic data, but also supports the principled comparison of augmentation techniques, especially important in applications where preserving subtle emotional and stylistic cues is crucial, such as in affective computing or mental health analysis.

### 2.6 Research Gaps and Opportunities

Despite recent progress in emotion intensity classification and the application of generative AI, several significant research gaps remain.

#### 2.6.1 Limited availability of high-quality emotion intensity datasets

Emotion intensity is inherently subjective and subtle, making consistent annotation a challenging task. Most existing datasets are small in scale and suffer from class imbalance, particularly underrepresentation of high-intensity emotional states. Furthermore, there is a lack of cross-domain datasets that reflect the diversity of emotional expression across contexts (e.g., social media vs. therapeutic dialogue).

#### 2.6.2 Lack of augmentation methods tailored to emotional nuance

While generative models such as LLMs have demonstrated great potential for text augmentation, few studies have investigated whether these models can generate emotionally coherent and intensity-preserving samples. Existing augmentation techniques often prioritize fluency and diversity over emotional fidelity, leading to potential misalignment between synthetic texts and their intended emotion labels.

#### 2.6.3 Limited exploration of domain adaptation in affective computing

Cross-dataset generalization remains a challenge, particularly when transferring knowledge between datasets with different styles, vocabularies, or intensity scales. The potential of generative AI to bridge such gaps through domain-aware augmentation has not been sufficiently explored, and few frameworks offer mechanisms for aligning emotional expression styles across corpora.

## 3 Data Augmentation Methodology

To address the inherent challenges of limited labeled data in emotion intensity classification tasks, we developed a comprehensive data augmentation framework that leverages cross-dataset knowledge transfer between target dataset (ESConv [Liu et al., 2021]) and source dataset (MEISD [Firdaus et al., 2020]). This approach combines traditional EDA techniques with modern large language model capabilities to generate high-quality synthetic training samples while preserving the linguistic and emotional characteristics of the target domain.

The framework operates on the principle that effective data augmentation for emotion intensity classification requires not only increasing the quantity of training data but also ensuring that the augmented samples maintain the stylistic and semantic properties of authentic emotional support conversations. Rather than applying generic text augmentation techniques, our methodology incorporates domain-specific knowledge extracted from the target dataset to guide the transformation process, ensuring that original samples are adapted to match the conversational patterns and emotional expression styles characteristic of the target domain.

This framework not only increases data availability but also prioritizes semantic integrity and class balance—two critical aspects often missing from small, imbalanced emotion datasets. By integrating synthetic generation with class-aware balancing, it helps mitigate data scarcity and skewed distribution problems common in affective tasks.

### 3.1 Intensity Label Mapping and Preprocessing

The foundation of this augmentation approach lies in establishing consistent intensity representations across different datasets, each originally annotated with distinct labeling schemes. This preprocessing step is crucial for ensuring that the augmented samples maintain meaningful emotional intensity distinctions while enabling effective cross-dataset knowledge transfer.

Two complementary corpora were used to enable sequential transfer learning between domains of emotional expression. The MEISD dataset [Firdaus et al., 2020] comprises scripted TV dialogues annotated on a 1–3 intensity scale, while ESConv [Liu et al., 2021] contains authentic emotional-support conversations rated on a 1–5 self-reported scale. To unify these data within a binary classification framework, scores ≤ 1.5 in MEISD were mapped to low intensity (0) and *>* 1.5 to high intensity (1); for ESConv, labels 1–2 were mapped to low (0) and 3–5 to high (1). This normalization ensured comparable intensity distributions and enabled two-phase training, where the model was first pretrained on MEISD (source domain) and subsequently fine-tuned on ESConv (domain-specific). Detailed dataset statistics and annotation examples are provided in Table 3.1.

**Table 3.1:** Comparison of source and target domains in the transfer learning framework, highlighting domain type, annotation style, and their role in model training.

| Aspect | Source domain | Target domain |
| --- | --- | --- |
| Domain | TV series dialogues (scripted, acted) | Emotional support conversations |
| Annotation type | External annotators | Self-reported intensity |
| Role in transfer | Source domain for pre-training | Target domain for fine-tuning |

**Table 3.2:** Structure of the LLM prompt used for intensity-conditioned message transformation, incorporating domain-specific stylistic cues and intensity-dependent generation instructions.

| Prompt Component | Description / Content |
| --- | --- |
| **Purpose** | Guide the large language model (LLM) to transform input messages so that they align stylistically and emotionally with real emotional support conversations, while maintaining semantic coherence and authenticity. |
| **Examples** | Each prompt includes three authentic examples from the target dataset representing the desired emotional intensity level. These examples illustrate how real users express emotions at that intensity, providing concrete references for tone, phrasing, and conversational structure.<br>*Prompt fragment:*<br>“You are helping transform messages to match the style of real emotional support conversations. Here are examples of [target intensity] messages from actual support conversations:” |
| **Key Stylistic Features** | Extracted automatically using the domain-specific style analysis module described in Section 3.2.1. Each prompt embeds concise stylistic cues characteristic of the target domain and intensity level, including:<br>• Common lexical patterns and keywords,<br>• Typical sentence length,<br>• Characteristic personal pronoun. |
| **Intensity-specific Instructions** | **Low intensity:**<br>“Transform this into a calm, reflective message that someone might share when feeling mildly concerned or seeking gentle support. The tone should be measured and thoughtful, focusing on understanding rather than expressing crisis.”<br>**High intensity:**<br>“Transform this into a message expressing significant emotional distress that someone might share when feeling overwhelmed and in urgent need of support. The tone should convey genuine emotional pain, immediacy, and a strong appeal for help.” |

**Table 3.3:**
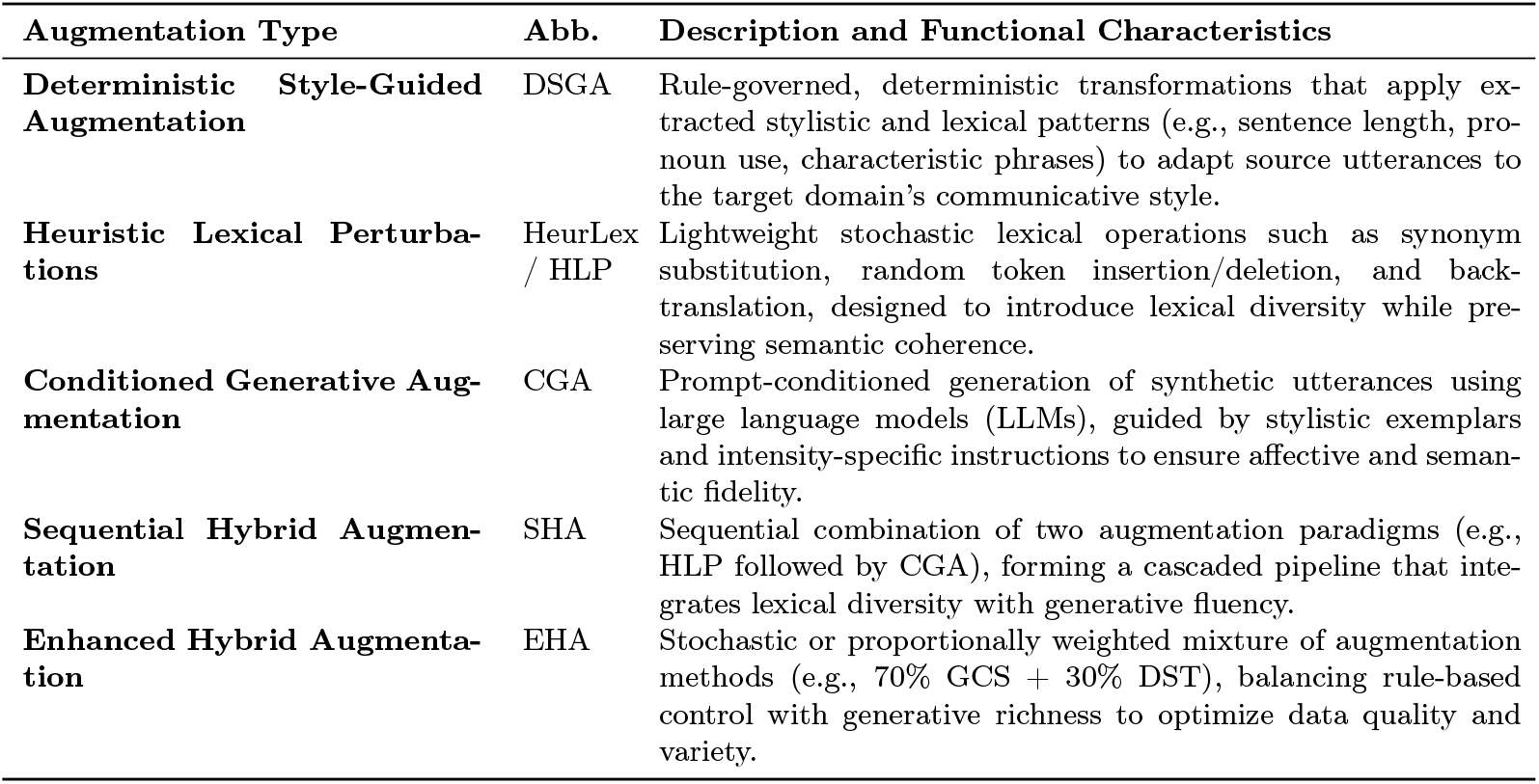
Summary of Data Augmentation Techniques and Their Functional Characteristics.

**Table 3.4:** Taxonomy of evaluation metrics applied within the quality assessment framework.

| Category | Metrics Used | Analytical Purpose |
| --- | --- | --- |
| Machine Translation Quality | BLEU | Surface-level lexical overlap between augmented and reference samples. Indicates paraphrasing precision. |
| Character-Level Similarity | CHRF | Morphology-sensitive similarity capturing fine-grained spelling and word form preservation. |
| Text Generation Quality | Self-BLEU, TTR, UTR, RWORDS, Novelty | Measures intra-set diversity, lexical variety, and novelty of generated samples. |
| Fluency and Grammaticality | Perplexity, Spelling Error Ratio | Estimates linguistic fluency and correctness of augmented samples. |
| Task-Specific Classification Quality | Accuracy, Precision, Recall, F1, AUC, Jaccard | Evaluates downstream model performance and robustness across augmentation types. |
| Semantic and Embedding Similarity | Cosine Similarity, Spearman Correlation | Quantifies dataset-level semantic proximity in embedding space. |

### 3.2 Stylistic Pattern Analysis

The core innovation of our augmentation approach lies in the comprehensive analysis of stylistic patterns that characterize emotional support conversations. This analysis forms the foundation for all subsequent transformation processes, ensuring that augmented samples not only maintain semantic coherence but also exhibit the linguistic characteristics that define authentic emotional support exchanges.

#### 3.2.1 Target Domain Style Characterization

To ensure that augmented samples maintain the linguistic characteristics of the target domain, we implemented a comprehensive style analysis module that extracts domain-specific patterns across multiple linguistic dimensions. This analysis operates on the principle that effective emotional support conversations exhibit distinctive patterns in vocabulary usage, sentence structure, and emotional expression that must be preserved in augmented samples.

*Lexical feature analysis* employs n-gram extraction [Cavnar and Trenkle, 1994] to identify common sentence starters, recognizing that the way individuals begin their emotional expressions often reflects the intensity and nature of their emotional state. Term Frequency-Inverse Document Frequency (TF-IDF) vectorization [Salton and Buckley, 1988] is applied to identify intensity-specific keywords, creating vocabulary profiles that characterize different levels of emotional intensity. Personal pronoun usage patterns are analyzed to capture the self-referential nature of emotional support seeking, with particular attention to first-person pronouns that indicate personal disclosure and emotional vulnerability.

*Syntactic feature analysis* focuses on structural elements that distinguish different intensity levels, including average sentence length calculations per intensity level and the frequency of questions and exclamations. These features capture the relationship between emotional intensity and linguistic expression patterns, with higher intensity conversations often exhibiting shorter, more fragmented sentences and increased use of punctuation that conveys emotional emphasis.

*Semantic feature analysis* employs TF-IDF vectorization with n-gram ranges of 1-2 to extract intensity-specific vocabulary, identifying the top-20 characteristic phrases for each intensity level. This analysis reveals the specific language patterns that distinguish high-intensity from low-intensity emotional expressions, providing concrete targets for transformation processes.

Such detailed linguistic profiling ensures that augmented samples are emotionally plausible and contextually faithful to the support-seeking tone of real interactions, rather than merely grammatically correct. This enables the transformation pipeline to produce not just fluent, but affectively aligned outputs.

#### 3.2.2 Cross-Dataset Pattern Comparison

The framework performs comparative analysis between target and source datasets linguistic patterns to identify specific transformation targets and guide the augmentation process. This comparative approach recognizes that effective domain adaptation requires understanding not only the characteristics of the target domain but also the specific differences between source and target domains that must be addressed through transformation.

The analysis includes distribution analysis of sentence lengths across intensity levels, revealing how emotional intensity correlates with linguistic expression patterns in each dataset. Vocabulary overlap assessment between datasets identifies shared and unique linguistic elements, informing decisions about which aspects of the source text should be preserved versus transformed. Stylistic gap identification provides specific targets for transformation, highlighting the linguistic features that most strongly distinguish the two datasets and require attention during the augmentation process.

### 3.3 Enhanced Transformation Methods

#### 3.3.1 Deterministic Style-Guided Augmentation

Following the style characterization phase, the classical transformation stage applies the extracted linguistic patterns to systematically realign source samples with the emotional and structural conventions observed in target dataset. This deterministic style-guided procedure translates the quantitative stylistic insights from the domain analysis into deterministic linguistic modifications, ensuring that each transformed utterance reflects the characteristic emotional intensity and discourse style of real support-seeking interactions.

For low-intensity transformations, the process attenuates affective tone through the replacement of highly charged lexical items with semantically equivalent but emotionally neutral expressions, and through the insertion of reflective linguistic cues that convey calmness and self-awareness. Conversely, high-intensity transformations enhance expressive salience by introducing intensity-specific lexical markers identified in the target corpus and by emphasizing first-person constructions that increase immediacy and emotional resonance.

To preserve stylistic authenticity, syntactic structure and sentence length are further adjusted toward the statistical norms of the target intensity level, mirroring the pacing and rhythm characteristic of natural emotional disclosure. The resulting utterances maintain the semantic coherence of the original input while exhibiting affective and structural properties empirically aligned with genuine emotional support discourse.

#### 3.3.2 Heuristic Lexical Augmentation

The heuristic lexical augmentation (HLA) strategy applies lightweight, rule-based manipulations to the original messages without leveraging pretrained language models. A technique is randomly selected from the following set: synonym replacement, random token insertion, random token deletion, and backtranslation. These modifications introduce lexical variation while preserving the core semantic content. This EDA-based heuristic augmentation extends standard EDA by incorporating domain-specific stylistic rules derived from the target dataset, ensuring transformed messages capture the conversational and emotional patterns characteristic of the target domain.

To improve domain consistency, each transformed message is further post-processed using target-domain stylistic adaptation rules. These include adjustments based on expected word count, use of personal pronouns, and intensity-specific phrasing patterns. This ensures that even purely heuristic augmentations reflect the stylistic signature of the target emotional domain.

Although this approach lacks the semantic fluency of LLM-based generation, it offers a low-resource, interpretable baseline for affective data augmentation, and serves as a valuable contrast in assessing the importance of semantic richness and emotional fidelity.

#### 3.3.3 Generative Conditioned Augmentation

The generative conditioned augmentation (GCA) method leverages a few-shot, in-context prompting strategy, combining exemplar-based and instruction-guided prompts to generate target-style utterances. This implementation employs a carefully designed prompting strategy that integrates domain-specific knowledge extracted from the target dataset to guide the transformation process. The fundamental principle underlying this approach is that effective domain adaptation requires not only semantic understanding but also stylistic consistency with the target domain.

The prompt engineering process begins with the dynamic construction of intensity-specific prompts that incorporate multiple layers of contextual information. For each transformation, the system selects 3 authentic examples from the target dataset as in-context demonstrations, guiding the LLM to produce outputs aligned with the desired emotional intensity and conversational style. These examples are complemented by extracted stylistic characteristics, including average sentence length, characteristic vocabulary, and personal pronoun usage patterns specific to each intensity level. This contextualized prompting approach ensures that the model has sufficient information to produce transformations that align with the linguistic patterns observed in real emotional support conversations.

The transformation instructions are carefully crafted to capture the nuanced differences between low and high intensity emotional expressions. For *low intensity transformations*, the system instructs the model to generate calm, reflective messages that convey mild concern while maintaining a measured tone focused on seeking understanding rather than expressing crisis. This approach recognizes that low-intensity emotional support seeking often involves thoughtful self-reflection and gentle requests for guidance. Conversely, *high intensity transformations* are guided by instructions that emphasize the expression of significant emotional distress, conveying genuine emotional pain and urgency for help while maintaining the authentic voice of someone experiencing overwhelming feelings (Table **??**). The transformation process was therefore designed not only for surface-level rewriting, but to embed affective intensity and interpersonal tone directly into the output, making it more suitable for emotion-sensitive applications.

The technical implementation utilizes the LLaMA-2-7B-Chat model [Touvron et al., 2023b] with 4-bit quantization, balancing computational efficiency with transformation quality. The context window of 2048 tokens provides sufficient space for comprehensive prompts while maintaining reasonable processing times. The temperature parameter of 0.7 was selected to encourage creative variation while preventing excessive randomness that could compromise the quality of transformations. Output generation is constrained to 200 tokens maximum per transformation, ensuring that the generated content remains focused and relevant while matching the typical length patterns observed in the target dataset.

**Figure 3.1:**
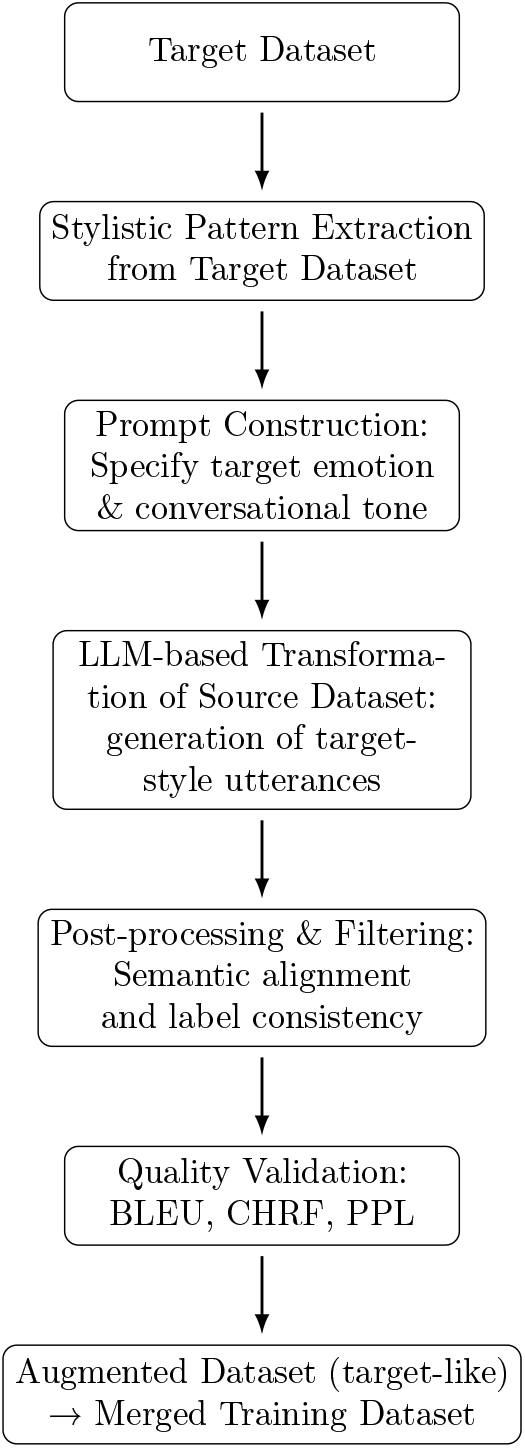
LLM-based augmentation pipeline converting source samples into target-style utterances for cross-domain adaptation.

#### 3.3.4 Sequential Hybrid Augmentation

The sequential hybrid augmentation (SHA) method combines both non-neural and neural approaches by sequentially or conditionally applying heuristic lexical perturbation (described in Section 3.3.2) and conditioned generative transformations (Section 3.3.3). In each run, the system randomly decides whether to begin with the CGA or HLA. If the initial transformation succeeds, the resulting message is refined using the complementary method. For instance, an LLM-generated output may be passed through a synonym replacement module, or a heuristically modified input may be rewritten via the LLM prompt described in Section 3.3.3.

This strategy is intended to test whether hybridization can combine the structural variation introduced by lexically-driven edits with the semantic fluency and emotional coherence enabled by large language models. It also simulates realistic scenarios where augmentation must operate under varying resource constraints or pipeline configurations.

#### 3.3.5 Enhanced Hybrid Augmentation

The proposed enhanced hybrid augmentation (EHA) method extends the baseline hybrid approach by combining rule-based and neural transformations in a dynamically weighted manner. Approximately 70% of samples are generated through LLM-based rewriting (Section 3.3.3), while 30% use heuristic lexical perturbations (Section 3.3.2). This balance enhances both semantic richness and stylistic fidelity.

To ensure domain consistency, EHA integrates stylistic features extracted from the target dataset, such as average sentence length, pronoun frequency, and intensity-specific vocabulary. These parameters guide both LLM prompt construction and heuristic perturbations, aligning outputs with the linguistic characteristics of emotional support conversations.

Each transformation follows a conditional workflow: if the initial operation yields a coherent output, it is refined using the complementary method. For instance, LLM-generated text may undergo synonym replacement, or rule-perturbed sentences may be recontextualized via prompt-based rewriting. The resulting data are validated for semantic consistency and stylistic quality using BLEU, CHRF, and perplexity metrics, forming a balanced, domain-adapted augmented corpus.

### 3.4 Quality Assessment Framework

To ensure comprehensive evaluation of our augmentation methods, we developed a multi-layered quality assessment framework that extends beyond traditional lexical similarity measures. This framework integrates established NLP evaluation metrics with domain-specific transformation quality and stylistic consistency measures, providing a holistic view of augmentation quality across linguistic, semantic, and emotional dimensions.

#### 3.4.1 Transformation Quality Assessment

Our primary quality evaluation mechanism employed a transformation quality scoring system that assessed the effectiveness of each augmentation method in generating high-quality synthetic samples. Each augmented sample was evaluated against the most similar original sample from the source dataset, with quality scores computed based on semantic coherence, structural integrity, and emotional appropriateness.

The quality of each transformed utterance was evaluated using a composite quality score that integrates three criteria: length similarity, keyword presence, and personal pronoun usage. Let *L*_*t*_ be the length (in words) of the transformed utterance, and *L*_*target*_ be the average length for the target intensity class in the domain-specific dataset. The length score is defined as

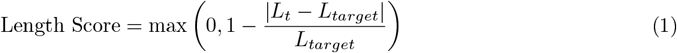

Let *K*_*m*_ denote the number of intensity-specific keywords (up to 10) appearing in the transformed text. The **keyword score** is given by

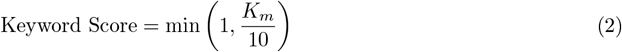

Finally, the **personal pronoun score** is

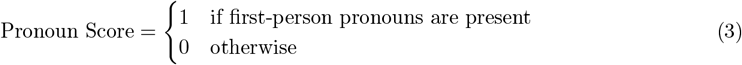

The overall quality score is computed as a weighted sum:

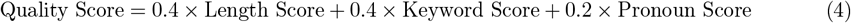

This metric captures both stylistic alignment with the target intensity class and the semantic incorporation of relevant emotional cues.

**Average Quality Score** represents the mean transformation quality across all augmented samples:

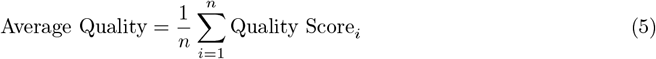

where *n* is the total number of augmented samples.

**Quality Range** captures the span of transformation quality variation:

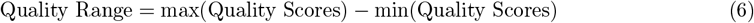

**Uniqueness Ratio** measures the proportion of unique texts in the augmented dataset:

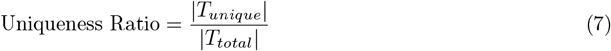

where *T*_*unique*_ represents the set of unique conversation texts and *T*_*total*_ is the complete set of generated texts.

**Average Text Length** quantifies the mean word count across augmented samples:

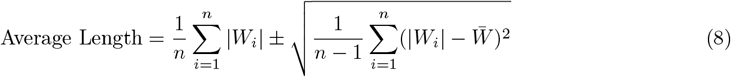

where |*W*_*i*_| represents the word count of text and the second term indicates the standard deviation.

**Balance Ratio** ensures equal class distribution in balanced datasets:

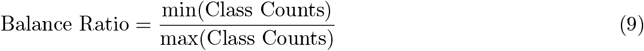

A ratio of 1.0 indicates perfect class balance.

#### 3.4.2 Stylistic Consistency Evaluation

The stylistic consistency assessment measured how well augmented samples preserved the characteristic linguistic patterns and stylistic elements of the target domain. This evaluation was conducted separately for low-intensity and high-intensity emotional expressions, recognizing that different emotional intensities exhibit distinct stylistic characteristics.

This framework analyzed multiple stylistic dimensions including sentence length patterns, vocabulary choices, personal pronoun usage, and conversation starter preferences. The consistency scores were computed by comparing augmented samples against established style patterns extracted from the targeted dataset.

**Low Intensity Consistency** was computed by averaging the stylistic similarity scores between augmented text samples (*T*_*i*_) and low-intensity reference patterns (*P*_*low*_) across all samples classified as low intensity:

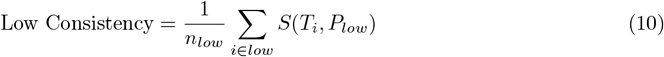

where *n*_*low*_ represents the total number of low-intensity samples, and the summation occurs over all samples *i* ∈ *low*.

**High Intensity Consistency** followed an analogous approach for high-intensity emotional expressions:

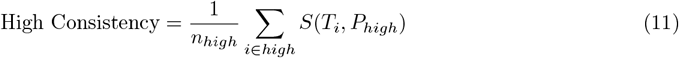

where *n*_*high*_ represents the total number of high-intensity samples, and the summation occurs over all samples *i* ∈ *high*.

The **Average Consistency** was calculated as the arithmetic mean of the intensity-specific consistency scores:

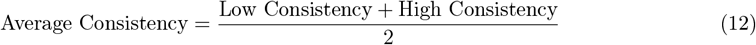

This approach ensures equal weighting between low and high-intensity stylistic patterns, regardless of the distribution of samples across intensity levels in the dataset.

##### Stylistic Similarity Score

The core of the evaluation framework relied on a composite stylistic similarity score *S*(*T*_*i*_, *P*_*j*_) that quantified the alignment between a given text sample *T*_*i*_ and reference patterns *P*_*j*_ across multiple linguistic dimensions:

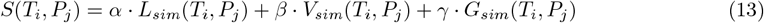

where the weighting coefficients satisfy the constraint *α* + *β* + *γ* = 1.

The three components of this similarity score capture distinct aspects of stylistic consistency:

- *L*_*sim*_(*T*_*i*_, *P*_*j*_): Lexical similarity measuring vocabulary choice alignment and word usage patterns
- *V*_*sim*_(*T*_*i*_, *P*_*j*_): Variability similarity assessing sentence structure and length distribution consistency
- *G*_*sim*_(*T*_*i*_, *P*_*j*_): Grammatical similarity evaluating syntactic patterns and grammatical structure preservation

The weighting parameters *α, β*, and *γ* allow for domain-specific calibration, enabling researchers to emphasize particular stylistic dimensions based on the characteristics most relevant to emotional support conversations. This weighted combination provides a comprehensive assessment of how well augmented samples maintain the distinctive stylistic fingerprint of the original target dataset across different emotional intensity levels.

#### 3.4.3 Standard NLP Evaluation Metrics

To complement the internal transformation and stylistic consistency evaluations, we conducted an additional layer of analysis based on widely adopted NLP evaluation metrics, as proposed in the unified taxonomy by Amadeus and Cruz Castañeda [2024]. These metrics provide insight into the linguistic quality, diversity, and fluency of augmented samples, enabling us to assess not only their alignment with the original data but also their utility for downstream modeling. The following metrics were computed for all balanced augmented sets:

**BLEU (Bilingual Evaluation Understudy)** [Papineni et al., 2002] evaluates n-gram overlap between generated samples and their source or reference counterparts. Although originally developed for machine translation, BLEU provides a surface-level measure of lexical similarity in data augmentation pipelines. Higher BLEU scores suggest close paraphrasing, while lower scores indicate more novel transformations.

The BLEU score is calculated as:

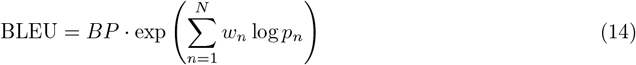

where *BP* represents the brevity penalty that penalizes sequences shorter than the reference, *w*_*n*_ denotes the weight assigned to each n-gram level (typically uniform with *w*_*n*_ = 1*/N*), and *p*_*n*_ is the precision score for n-grams of length n.

**CHRF (Character n-gram F-score)** [Popović, 2015] captures character-level similarity, offering a language-agnostic and morphology-sensitive alternative to BLEU. It is particularly useful in assessing whether paraphrases preserve linguistic structure and subtle word forms.

The CHRF score is computed as:

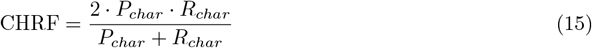

where *P*_*char*_ and *R*_*char*_ represent character-level precision and recall respectively, calculated as:

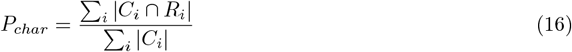

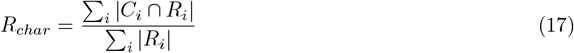

Here, *C*_*i*_ represents the set of character n-grams in the candidate text, *R*_*i*_ represents the character n-grams in the reference text, and |*C*_*i*_ ∩ *R*_*i*_| denotes their intersection.

**Self-BLEU** [Zhu et al., 2018] is a metric for measuring the internal diversity of generated samples. Each augmented sentence is treated as a hypothesis and compared to all others as references. High Self-BLEU indicates redundancy (low diversity), while lower values reflect greater variation—desirable in data augmentation contexts.

Self-BLEU is calculated as:

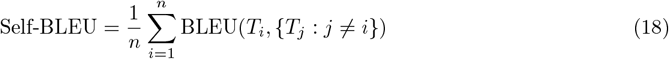

where *n* is the total number of generated samples, *T*_*i*_ is the i-th text treated as hypothesis, and {*T*_*j*_ : *j* ≠ *i*} represents all other texts serving as references.

**Perplexity (PPL)** is used to evaluate the fluency of generated samples by measuring how well a language model predicts them. Lower perplexity scores indicate that the text is more grammatically and syntactically plausible, resembling natural language. In our case, GPT-2 perplexity scores were used to estimate the fluency of each augmentation method.

Perplexity is calculated as:

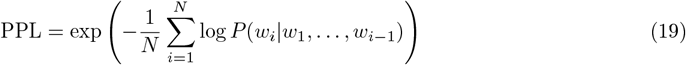

where *N* is the total number of tokens, *w*_*i*_ represents the i-th token, and *P* (*w*_*i*_|*w*_1_, …, *w*_*i*−1_) is the conditional probability of the i-th token given all preceding tokens.

##### Diversity Assessment Metrics

While the framework includes general-purpose metrics such as BLEU, CHRF, and Perplexity, it also considers diversity and stylistic consistency—reflecting the need for more nuanced evaluation criteria that go beyond lexical similarity. This hybrid evaluation approach moves toward filling the gap in emotion-aware quality assessment.

##### TTR (Type-Token Ratio)

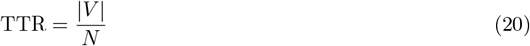

where |*V* | represents the vocabulary size (number of unique words) and *N* is the total number of tokens.

##### UTR (Unique Trigram Ratio)

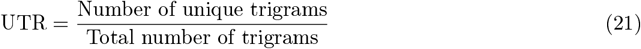

This metric measures the proportion of unique three-word sequences relative to all trigrams in the dataset.

##### RWORDS

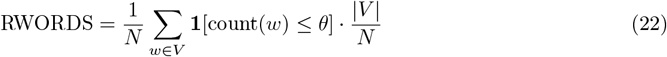

where *θ* is a threshold parameter, *N* is the total token count, count(*w*) represents the frequency of word *w*, and |*V*| is the vocabulary size.

##### Novelty Score

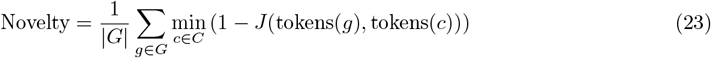

where *g* represents a generated sample, *C* is the set of original samples, and *J* (*A, B*) = |*A* ∩ *B*| */* |*A* ∪ *B*| represents the Jaccard similarity coefficient between token sets.

### 3.5 Data Augmentation Framework

The framework implements two distinct augmentation strategies designed to address different aspects of the data scarcity problem in emotion intensity classification. These strategies recognize that effective data augmentation may require different approaches depending on the specific characteristics of the original dataset and the intended application.

#### Proportional Augmentation

This strategy increases the dataset size by a predefined percentage while preserving the original class distribution. It is particularly effective when the dataset already exhibits an acceptable class balance, and the main objective is to expand the overall amount of training data without altering its statistical properties. By proportionally generating new samples for each class, the model can generalize better without introducing additional bias.

#### Balanced Augmentation

In contrast, the balanced augmentation strategy focuses on mitigating class imbalance by generating additional samples for underrepresented classes until all classes reach the same number of instances as the majority class. This method is especially useful when the dataset contains significant disparities between emotion intensity levels, as it helps ensure that the model receives an equal learning opportunity across all categories. As a result, it reduces bias toward dominant classes and can improve the robustness of classification performance on minority emotions.

### 3.6 Validation and Output

The final stage of the augmentation framework focuses on ensuring dataset integrity and providing comprehensive outputs that support subsequent analysis and model training.

Post-augmentation validation ensures that the final datasets meet quality standards and maintain the intended characteristics. Distribution analysis verifies that the augmented datasets achieve the intended class balance and that the augmentation process has not introduced unexpected biases or artifacts. Quality distribution analysis provides statistical summaries of transformation quality scores, enabling assessment of the overall effectiveness of different augmentation methods.

Duplication detection [Broder, 1997] identifies and removes near-duplicate samples that might have been generated through the augmentation process, ensuring that the final datasets contain diverse and unique training examples.

## 4 Classification Methodology

### 4.1 Model Architecture for Classification Problem

We developed a hybrid neural architecture that integrates BERT (Bidirectional Encoder Representations from Transformers) [Devlin et al., 2019] with Long Short-Term Memory (LSTM) networks [Hochreiter and Schmidhuber, 1997] for binary intensity classification tasks. The architecture employs a three-stage processing pipeline designed to leverage both contextual understanding and temporal sequence modeling capabilities.

The initial stage utilizes BERT-base-cased as the primary encoder, generating dense contextual embeddings for input sequences. We enhanced the model’s regularization through strategic implementation of dropout mechanisms [Srivastava et al., 2014], setting both hidden dropout probability and attention dropout probability to 0.3. This configuration was selected to balance model expressiveness with generalization capability, preventing overfitting while maintaining the transformer’s capacity for complex pattern recognition.

The intermediate stage processes BERT output sequences through a bidirectional LSTM layer [Hochreiter and Schmidhuber, 1997] configured with 64 hidden units in a single-layer architecture. The bidirectional design enables the model to capture temporal dependencies in both forward and backward directions, providing comprehensive sequence-level representations that complement the token-level embeddings from the transformer component.

The final classification stage consists of a two-layer fully connected network incorporating batch normalization [Ioffe and Szegedy, 2015] and dropout regularization. The hidden layer employs a dimension equal to half the LSTM output size, followed by a single-unit output layer with sigmoid activation. This design ensures stable gradient flow during training while producing well-calibrated probability estimates for binary classification decisions. The combination of batch normalization and dropout regularization provides robust training dynamics and improved generalization performance.

### 4.2 Training Strategy and Regularization

We implemented a comprehensive training strategy incorporating multiple regularization techniques to optimize model performance and prevent overfitting. The training process follows a two-phase approach to maximize learning efficiency across both datasets. To address overfitting and enhance generalization, we incorporated multiple regularization strategies during training, detailed in the following subsection.

The loss function utilized Focal Loss [Lin et al., 2017] as the primary objective to address potential class imbalance issues, with *α* = 2.0 and *γ* = 1.5 parameters. The optimization process employed the AdamW optimizer [Loshchilov and Hutter, 2019] with dataset-specific learning rates: 5e-6 for source dataset pretraining and 2e-6 for target dataset fine-tuning. Learning rate scheduling was implemented with patience of 3 epochs and reduction factor of 0.3 to adapt the learning rate based on validation performance. The complete training configuration, including dataset-specific hyperparameters and regularization settings, is summarized in Table 4.1.

**Table 4_1:** Training Configuration Summary.

| Parameter | Source Dataset<br>Pre-training | Target Dataset<br>Fine-tuning |
| --- | --- | --- |
| Max Epochs | 15 | 3 |
| Learning Rate | 5e-6 | 2e-6 |
| Batch Size | 16 | 16 |
| Early Stopping | 3 | 1 |
| Frozen Layers | 0-9 | 0-7 (8-11 unfrozen) |
| Dropout Rate | 0.3 (BERT),<br>0.5 (classifier) | 0.3 (BERT),<br>0.5 (classifier) |
| Weight Decay | 0.01 | 0.01 |

#### 4.2.1 Two-Phase Training Approach

As illustrated in Figure 4.1, the model follows a two-phase training process to optimize model performance across both datasets:

**Figure 4.1:**
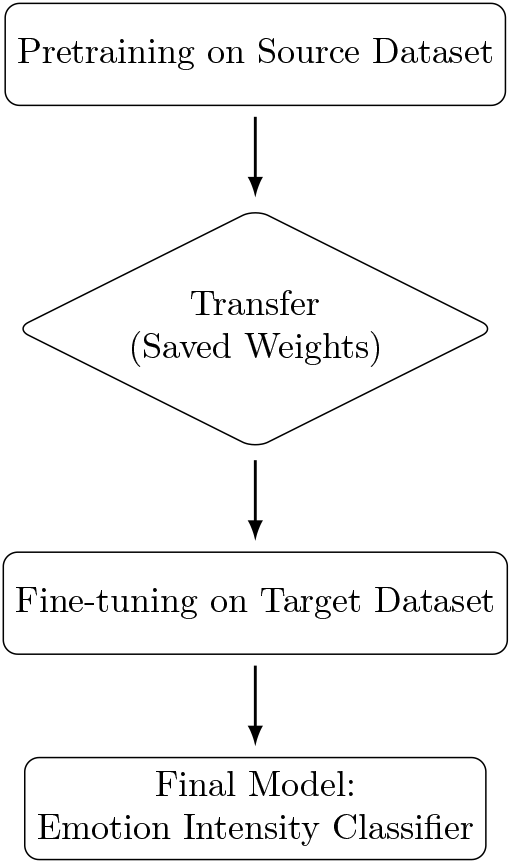
Overview of the twophase training workflow consisting of (1) pretraining on the source dataset and (2) fine-tuning on the target dataset using transferred weights. The resulting model serves as the final emotion intensity classifier.

##### Phase 1: Pre-training on Source Dataset

The model undergoes initial training on the source dataset for up to 15 epochs, establishing foundational representations for intensity classification. Early stopping [Prechelt, 1998] was implemented with patience of 3 epochs to prevent overfitting while ensuring adequate model convergence.

##### Phase 2: Fine-tuning on Target Dataset

The pre-trained model is subsequently fine-tuned on the target dataset for up to 3 epochs, adapting the learned representations to the target domain. During this phase, the last 4 layers of BERT (layers 8–11) were unfrozen to allow for domain-specific adaptation [Lee et al., 2021]. Early stopping was triggered with patience of 1 epoch or when validation accuracy reached 98% to prevent overfitting on the smaller fine-tuning dataset.

This sequential training paradigm allows the model to learn general emotional patterns from augmented data and later adapt to the nuances of the target domain. Such a setup is particularly valuable when training across stylistically divergent corpora, as it encourages domain bridging and controlled adaptation. Beyond its standalone predictive purpose, this two-phase classifier also serves as an analytical instrument for evaluating the effectiveness of various data augmentation strategies. Specifically, it enables quantifying how well different augmentation methods preserve or enhance the emotional intensity expressed in textual data.

#### 4.2.2 Regularization Techniques

To address overfitting concerns, we implemented multiple regularization strategies:

##### Layer Freezing

During initial training, we froze the first 10 layers of BERT (layers 0-9) along with the embedding layers, allowing only the upper layers to adapt to the task.

##### Dropout Regularization

Applied dropout with rates of 0.5 for the classification layers and 0.3 for BERT components.

##### Weight Decay

Implemented L2 regularization with a weight decay coefficient of 0.01.

##### Gradient Clipping

Applied gradient clipping with a maximum norm of 1.0 to prevent gradient explosion.

#### 4.2.3 Loss Function and Optimization

We employed Focal Loss [Lin et al., 2017] as the primary loss function to address potential class imbalance issues, with parameters *α* = 2.0 and *γ* = 1.5:

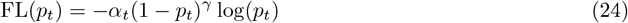

where *p*_*t*_ is the model’s estimated probability for the true class, *α*_*t*_ is a weighting factor for class *t*, and *γ* ≥ 0 is the focusing parameter that reduces the relative loss for well-classified examples.

The model was optimized using the AdamW optimizer [Loshchilov and Hutter, 2019]. An adaptive learning rate scheduler was applied to reduce the learning rate when validation performance plateaued.

### 4.3 Data Preprocessing, Tokenization and Implementation Details

Text preprocessing was performed using the BERT tokenizer [Devlin et al., 2019] with addition of special tokens ([CLS], [SEP]), truncation and padding to maximum sequence length of 100 tokens, and attention mask generation for variable-length sequences. The model training employed a batch size of 16 samples per batch to balance computational efficiency with gradient stability. We employed stratified train-validation splits [Kohavi, 1995] (80:20) to ensure balanced representation of both intensity classes across training and validation sets, with standard random splitting applied when stratification was not possible due to class distribution constraints.

Model performance was assessed using multiple evaluation metrics including overall classification accuracy, weighted F1-score to account for potential class imbalance, class-specific precision and recall measures, and comprehensive loss tracking for convergence analysis. The best-performing model for each phase was selected based on minimum validation loss, with additional monitoring of validation accuracy to ensure optimal performance. Model states were preserved at each epoch’s best performance, enabling recovery of optimal parameters for final evaluation.

## 5 Results

### 5.1 Data Augmentation Quality Assessment

#### 5.1.1 Transformation Quality Analysis

The comprehensive quality assessment of the five augmentation methods revealed significant differences in their ability to generate high-quality synthetic samples. The evaluation was conducted on augmented datasets containing 4,738 samples each, with perfect class balance (2,369 samples per class) achieved through balanced augmentation strategy (Section 3.5).

The Conditioned Generative Augmentation (CGA) method demonstrated superior performance across all quality metrics. The enhanced CGA approach achieved the highest average transformation quality score of 0.719 ± 0.161, with quality scores ranging from 0.107 to 0.999 (Table 5.1). This method also exhibited the highest text uniqueness ratio of 0.999, indicating minimal duplication in generated samples, and produced texts with an average length of 87.9 ± 56.6 words, closely matching the target domain characteristics.

**Table 5.1:** Summary of data augmentation quality metrics across five methods.

| Method | Total Samples | Balance Ratio | Uniqueness Ratio | Avg Text Length (words) | Avg Quality Score | Quality Range |
| --- | --- | --- | --- | --- | --- | --- |
| DSGA | 4,738 | 1.000 | 0.896 | 68.5 | $0.398 \pm 0.157$ | 0.103–0.801 |
| HLA | 4,738 | 1.000 | 0.975 | 67.4 | $0.328 \pm 0.086$ | 0.108–0.600 |
| CGA | 4,738 | 1.000 | 0.999 | 87.9 | $0.719 \pm 0.161$ | 0.107–0.999 |
| EHA | 4,738 | 1.000 | 0.988 | 82.0 | $0.623 \pm 0.215$ | 0.104–0.999 |
| SHP | 4,738 | 1.000 | 0.991 | 87.9 | $0.446 \pm 0.124$ | 0.180–0.600 |

The Enhanced Hybrid Augmentation, combining 70% CGA (see Section 3.3.3) and DSGA (see Section 3.3.1), achieved intermediate performance with an average quality score of 0.623±0.215 (Table 5.1). This method demonstrated a quality range from 0.104 to 0.999, with a uniqueness ratio of 0.988 and average text length of 82.0±60.7 words. The wider standard deviation in quality scores suggests greater variability in transformation effectiveness compared to the CGA approach.

The Sequential Hybrid method achieved 0.446±0.124 quality, combining heuristic techniques with LLM refinements. It produced highly unique samples (0.991) and relatively long texts (87.7±57.9 words), indicating partial stylistic and structural integration of LLM output.

The Heuristic Lexical approach (no LLMs involved) recorded the lowest transformation quality (0.328±0.086) and narrower quality range (0.108–0.600), though uniqueness remained high (0.975). It also generated the shortest samples (67.4±68.2 words) alongside the classical method.

The Deterministic Style-Guided Augmentation method showed the lowest overall performance, with an average quality score of 0.398 ± 0.157 and quality range from 0.103 to 0.801 (Table 5.1). This method produced the shortest texts on average (68.5 ± 67.5 words) and achieved the lowest uniqueness ratio of 0.896, indicating higher levels of duplication in generated samples.

#### 5.1.2 Stylistic Consistency Evaluation

The analysis of stylistic consistency with the target domain revealed consistent patterns across intensity levels (Table 5.2). Unexpectedly, the augmentation methods incorporating NLP techniques (HLA and SHP) achieved the highest stylistic alignment, with consistency scores of 0.669 for low intensity and 0.721 for high intensity, yielding an average of 0.695. The LLM-based approach (CGA) followed closely with an average stylistic score of 0.617, indicating that while LLMs captured semantics effectively, they were marginally outperformed by rule-based heuristics (HLA) in reproducing surface-level stylistic traits.

**Table 5.2:** Stylistic Consistency Analysis.

| Method | Low Intensity<br>Consistency | High Intensity<br>Consistency | Average<br>Consistency |
| --- | --- | --- | --- |
| DSGA | 0.542 | 0.561 | 0.552 |
| HLA | 0.669 | 0.721 | 0.695 |
| CGA | 0.620 | 0.613 | 0.617 |
| EHA | 0.597 | 0.598 | 0.598 |
| SHP | 0.669 | 0.721 | 0.695 |

**Table 5_3:** NLP Metrics for Evaluating Transformation Quality.

| Method | BLEU | Self-BLEU | UTR | TTR | RWORDS | Misspelled<br>Words Ratio | Misspelled<br>Chars Ratio | PPL |
| --- | --- | --- | --- | --- | --- | --- | --- | --- |
| DSGA | 0.9798 | 0.8671 | 0.3619 | 0.0591 | 0.9938 | 0.0026 | 0.0016 | 38.94 |
| HLA | 0.9687 | 0.6381 | 0.5667 | 0.0748 | 0.9950 | 0.0028 | 0.0017 | 47.53 |
| CGA | 0.9989 | 0.7989 | 0.3206 | 0.0251 | 0.9950 | 0.0029 | 0.0021 | 35.55 |
| EHA | 0.9936 | 0.7484 | 0.3686 | 0.0322 | 0.9948 | 0.0030 | 0.0022 | 35.30 |
| SHP | 0.9994 | 0.8314 | 0.2642 | 0.0231 | 0.9933 | 0.0017 | 0.0006 | 33.44 |

The mixed approach (EHA), which combines LLM and rule-based transformations, demonstrated balanced consistency with scores of 0.597 for both low and high intensity levels. The classical method (DSGA) showed the lowest consistency scores of 0.542 for low-intensity and 0.561 for high-intensity transformations, with an average of 0.552 (Table 5.2).

These results suggest that the LLM-based approach most effectively captured the linguistic patterns and stylistic characteristics of emotional support conversations, while the classical rule-based method struggled to maintain the nuanced stylistic elements that characterize authentic emotional expressions.

#### 5.1.3 NLP Evaluation Metrics (BLEU, CHRF, Self-BLEU, Perplexity)

To complement the quality assessment of synthetic data, an additional evaluation was conducted using standard natural language generation metrics. The analysis included BLEU, CHRF, Self-BLEU, Type-Token Ratio (TTR), Unique Trigram Ratio (UTR), and spelling error rates. All metrics were computed on five balanced augmented datasets, each containing 2,450 samples.

**BLEU (Bilingual Evaluation Understudy)**, which measures n-gram overlap between the original and generated texts, showed consistently high values across all methods. The highest scores were achieved by the SHP hybrid (0.9994) and CGA (0.9989) approaches, while the lowest was observed for the HLA method (0.9687).

**CHRF (Character n-gram F-score)**, which captures character-level similarity and is independent of tokenization, reached the maximum score of 1.0000 for all methods, suggesting that generated texts remain very close to the originals at the character level.

**Self-BLEU**, a metric used to assess intra-set redundancy, revealed the highest overlap (and thus lowest diversity) in the DSGA approach (0.8671) and EHA approach (0.8314). More diverse outputs were observed for the HLA method (0.6381) and the EHA (0.7484).

**Lexical diversity metrics** confirmed these patterns. The HLA method achieved the highest UTR (0.5667) and TTR (0.0748), indicating greater lexical variation. In contrast, the SHP showed the lowest UTR (0.2642) and TTR (0.0231), reflecting higher redundancy.

**Spelling metrics** showed very low error rates overall. The SHP method achieved the lowest ratio of misspelled characters (0.0006), while the highest error rate was found in the EHA (0.0022).

**Perplexity (PPL)**, which reflects the fluency and predictability of generated text, was lower for methods incorporating large language models. The SHP hybrid (33.44), CGA (35.55), and EHA (35.30) produced the most coherent outputs, whereas higher perplexity values for the non-LLM methods, HLP (47.53) and DSGA (38.94), indicating comparatively lower fluency.

The comparative analysis reveals clear distinctions in the linguistic quality of the augmented datasets. LLM-based approaches, including CGA and the SHP hybrid, achieve the highest BLEU scores and lowest perplexity, indicating superior fluency and semantic alignment. However, these methods also exhibit higher Self-BLEU and lower lexical diversity, reflecting greater redundancy in phrasing. In contrast, the heuristic HLA method demonstrates the broadest lexical variety but reduced alignment with source structures. The classical rule-based DSGA approach produces moderately fluent yet repetitive outputs. Overall, the results confirm that LLM-driven augmentation ensures fluent and semantically consistent text, whereas heuristic strategies promote greater lexical diversity. The complementary strengths observed suggest that hybrid approaches, such as EHA, can balance semantic fidelity with stylistic and lexical variation, offering a promising direction for domain-adaptive text augmentation.

### 5.2 Classification Performance Results

#### 5.2.1 Overall Classification Performance Across Augmentation Strategies

Across both source and target datasets, augmentation had a strong positive impact on classification outcomes compared to models trained on the original, non-augmented data. Tables 5.4 and 5.5 summarize the core classification results for each method and dataset. Notably, the LLM-based augmentation (CGA) yielded the highest overall performance, particularly on original, achieving an F1 score of 0.8816, with a 95% confidence interval of [0.8610–0.9022], and an accuracy of 0.8819.

**Table 5_4:** Performance Summary of Augmentation Methods on Original Dataset.

| Augmentation | Accuracy | Precision | Recall | F1 Score | F1 Score CI |
| --- | --- | --- | --- | --- | --- |
| DSGA | 0.7395 | 0.74 | 0.7384 | 0.7392 | [0.6978, 0.7767] |
| HLA | 0.6593 | 0.6438 | 0.7131 | 0.6767 | [0.6345, 0.7161] |
| **CGA** | **0.8819** | **0.8835** | **0.8797** | **0.8816** | **[0.8494, 0.9077]** |
| **EHA** | **0.8323** | **0.8208** | **0.8502** | **0.8352** | **[0.7995, 0.8656]** |
| SHP | 0.7194 | 0.6948 | 0.7827 | 0.7361 | [0.6961, 0.7726] |
| Original dataset | 0.6498 | 0.7321 | 0.7987 | 0.7640 | [0.6928, 0.8228] |

**Table 5_5:** Performance Summary of Augmentation Methods on Target Dataset.

| Augmentation | Accuracy | Precision | Recall | F1 Score | F1 Score CI |
| --- | --- | --- | --- | --- | --- |
| **DSGA** | **0.8184** | **0.75** | **0.8673** | **0.8044** | **[0.7485, 0.8501]** |
| HLA | 0.7592 | 0.6809 | 0.8294 | 0.7479 | [0.6890, 0.7985] |
| **CGA** | **0.8204** | **0.7375** | **0.9052** | **0.8128** | **[0.7592, 0.8560]** |
| **EHA** | **0.8143** | **0.7439** | **0.8673** | **0.8009** | **[0.7449, 0.8468]** |
| SHP | 0.7959 | 0.711 | 0.8863 | 0.7890 | [0.7337, 0.8348] |
| Original dataset | 0.6918 | 0.6163 | 0.7536 | 0.6780 | [0.6161, 0.7342] |

The confidence intervals for F1-scores were calculated using the Wilson indirect method, which computes Wilson score intervals for precision and recall separately and derives F1-score bounds through interval arithmetic. This approach was selected based on recommendations by Takahashi et al. [2022], who demonstrated that Wilson-based methods show superior coverage probabilities compared to the Wald method, particularly when sample sizes are moderate. The Wilson indirect method is theoretically more rigorous than treating F1-scores as simple proportions, as it properly accounts for the harmonic nature of the F1 metric and the interdependence between precision and recall components. This methodological choice aligns with the broader literature advocating for score-based confidence interval construction over Wald-based approximations [Brown et al., 2001, Lam et al., 2023].

The EHA augmentation method also performed well on source dataset (F1 = 0.8352), significantly outperforming the DSGA method (F1 = 0.7392).

Interestingly, for the target dataset, although LLM augmentation (CGA) again reached the highest F1 score (0.8128), the differences among the top three augmentation strategies (CGA, DSGA, and EHA) were marginal (within ∼0.01 F1 points). This suggests that while augmentation improves performance overall, its relative benefit may diminish during transfer learning, especially when fine-tuning on a sufficiently large or stylistically aligned dataset like domain-specific dataset.

By contrast, HLA augmentation consistently underperformed in both datasets (F1 = 0.6767 for original dataset, 0.7479 for target dataset), and was even outperformed by the original original data (F1 = 0.7640). This suggests that while NLP methods offer linguistic diversity, they may not sufficiently capture the subtle semantic cues needed for high/low emotional intensity differentiation in this task.

#### 5.2.2 Statistical Significance and Effect Sizes for Accuracy

To further validate the observed performance differences, two-proportion z-tests were conducted on accuracy scores and computed Cohen’s *h* to quantify effect sizes.

For the original dataset (Table 5.6), statistically significant improvements were observed between:

**Table 5_6:** Statistical Comparisons of Accuracy on Original Dataset (Two-Proportion z-tests). Legend: *p* < 0.05 → *, *p* < 0.01 → **, *p* < 0.001 → ***.

|  | DSGA | HLA | CGA | EHA | SHP |
| --- | --- | --- | --- | --- | --- |
| **DSGA** | – | $z = 2.250$,<br>$p = 0.0244^*$ | $z = -3.762$,<br>$p = 0.0002^{***}$ | $z = -2.483$,<br>$p = 0.013^*$ | $z = 0.553$,<br>$p = 0.5804$ |
| **HLA** | $z = -2.250$,<br>$p = 0.0244^*$ | – | $z = 6.002$,<br>$p < 0.001^{***}$ | $z = 4.728$,<br>$p < 0.001^{***}$ | $z = -1.698$,<br>$p = 0.0895$ |
| **CGA** | $z = 3.762$,<br>$p = 0.0002^{***}$ | $z = -6.002$,<br>$p < 0.001^{***}$ | – | $z = -1.281$,<br>$p = 0.2002$ | $z = 4.313$,<br>$p = 0.0000^{***}$ |
| **EHA** | $z = 2.483$,<br>$p = 0.013^*$ | $z = -4.728$,<br>$p < 0.001^{***}$ | $z = 1.281$,<br>$p = 0.2002$ | – | $z = 3.035$,<br>$p = 0.0024^{**}$ |
| **SHP** | $z = -0.553$,<br>$p = 0.5804$ | $z = 1.698$,<br>$p = 0.0895$ | $z = -4.313$,<br>$p = 0.0000^{***}$ | $z = -3.035$,<br>$p = 0.0024^{**}$ | – |

- CGA vs. DSGA: *z* = −3.76, *p* = 0.0002, (acc: 0.1480 vs 0.1764)
- HLA vs. EHA: *z* = 4.73, *p* < 0.0001, (acc: 0.1665 vs 0.1319)
- CGA vs HLA: *z* = −6.00, *p* < 0.0001, (acc: 0.1764 vs 0.1319), with a Cohen’s *h* = 0.545, indicating a medium effect size
- CGA vs SHP: *z* = 4.313, *p* = 0.0000, (acc: 0.1764 vs 0.1439)

These results confirm that LLM-based augmentation significantly outperforms NLP methods and provides meaningful performance improvements over classical augmentation techniques.

For the target dataset (Table 5.7), however, no comparisons yielded statistically significant differences. This may reflect two key dynamics: (1) performance plateaus due to transfer learning from original and (2) greater variability in the target dataset, which can mask fine-grained improvements. Importantly, despite the lack of statistical significance, CGA still achieved the highest F1 score on target dataset (0.8128), with the narrowest confidence interval among all methods.

**Table 5_7:**
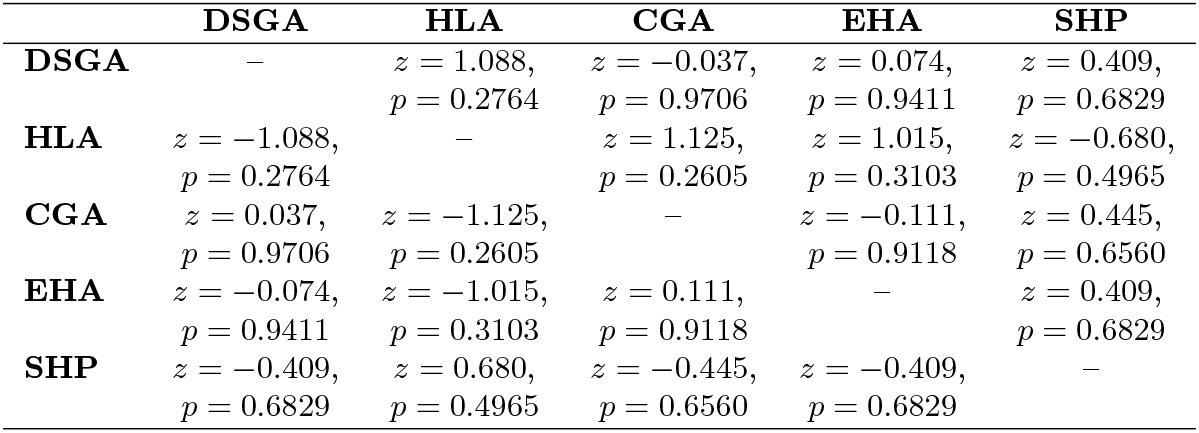
Statistical Comparisons of Accuracy on Target Dataset (Two-Proportion z-tests)

**Table 5_8:** Pearson Correlation Coefficients Between Augmentation Quality Metrics and F1 Classification Scores on the Original and Target Test Sets. Positive values indicate alignment between higher augmentation quality and better classification performance.

| Metric | Correlation with F1<br>(Original Dataset) | Correlation with F1<br>(Target Dataset) |
| --- | --- | --- |
| BLEU | 0.9986 | 0.802 |
| CHRF | 0.985 | 0.761 |
| Perplexity | -0.655 | -0.818 |
| Self-BLEU | 0.308 | 0.658 |

**Table 5_9:** Misclassified Examples (Qualitative Error Analysis)

| Case | Label | Pred. | Conf. | Excerpt | Interpretation |
| --- | --- | --- | --- | --- | --- |
| High-confidence but wrong prediction | Low | High | 0.948 | “Hi, i’ve been feeling really depressed lately and I’d like to talk to someone about it It’s a bit of everything. I’ve recently lost my job and I’m just really worried about how my bills are going to get paid. I feel so tired, you know? [...] I have no energy even to be thankful for the other little stuff [...] I could sleep forever but with my kids to worry about I don’t have that option [...] the little terrors are the reason I get up in the morning. I can’t let them down, you know” | This case is notable because the model was almost certain (>94% confidence) yet incorrect. It highlights a key weakness: insufficient separation between emotional *valence* and *intensity*. The model appears to conflate the presence of negative emotional states (e.g., *depressed*, *tired*, *worried*) with high emotional intensity, disregarding the calm, reflective tone of the speaker. |
| Ambiguous, low-confidence prediction | High | Low | 0.018 | “Yeah, thanks. I just hope it works out... Yeah, I’m using Indeed. Thanks, have you found a job? That’s true. It can be supplemented with gigs. Thanks for your help.” | This illustrates the intrinsic difficulty of the task. It represents a borderline case rather than a clear failure. The dialogue is short, polite, and lacks overt emotional markers. Contextual information beyond the text fragment (e.g., previous conversation turns) is required. |
| Disputed label with moderate confidence | Low | High | 0.521 | “Hi. Quite frustrated with my tween daughter! She doesn’t listen to anything I say. [...] I guess so. I’m 53 so going through menopause while she is going through puberty. True. I just wish she would take some pride in her things and herself. She hasn’t showered in a week and hates to brush her hair. [...] I have spoiled her rotten so that she has most everything she could want [...] She is getting much better about putting clean underwear on everyday at least.” | This case raises the possibility of annotation noise: the *Low* label may not accurately reflect the emotional intensity perceived by human annotators or the model during data augmentation process. The speaker expresses repeated frustration, annoyance, and exasperation through vivid examples. These linguistic cues align with a “High” intensity classification. |

Cross-dataset comparisons also reveal consistently significant *p*-values (*p* < 0.0001) when contrasting performance between original and target datasets for the same augmentation method, confirming that transfer learning had a measurable impact—models benefited from pretraining on augmented original before adapting to target dataset.

#### 5.2.3 Confidence Intervals and Robustness for F-score

The performance of various data augmentation methods on the original and target datasets is summarized in Tables 5.4 and 5.5, respectively, reporting accuracy, precision, recall, F1 Score, and 95% confidence intervals (CIs) for F1 Scores computed using the Confidence Indirect (CI) Wilson method [Brown et al., 2001, Lam et al., 2023]. While this method provides an approximate measure of uncertainty, it is important to note that F1 Score is a derived metric combining precision and recall, and thus the CIs may not fully capture its true variance.

On the original dataset, CGA augmentation achieved the highest F1 Score (0.8816) with a narrow confidence interval [0.8494, 0.9077], indicating high performance and stability. The combination of LLM and rule-based augmentation (EHA) also performed strongly (F1 = 0.8352, CI = [0.7995, 0.8656]), significantly outperforming rule-based DSGA augmentation alone (F1 = 0.7392, CI = [0.6978, 0.7767]).

HLA augmentation yielded the lowest F1 Score (0.6767) with a confidence interval of [0.6345, 0.7161], reflecting consistently weaker performance rather than random variability. Combining LLM with NLP in SHP pipeline (F1 = 0.7361, CI = [0.6961, 0.7726]) improved results relative to NLP alone (HLA method), suggesting a synergistic effect when multiple augmentation strategies are combined.

On the target dataset, CGA again achieved the highest F1 Score (0.8128, CI = [0.7592, 0.8560]), demonstrating robust performance across datasets. Those methods with rule-based DSGA and EHA augmentations produced similar F1 Scores (0.8044 and 0.8009, respectively) with overlapping confidence intervals, indicating no statistically significant difference between these methods on this dataset.

The original, non-augmented dataset consistently yielded the lowest F1 Score (0.678, CI = [0.6161, 0.7342]), showing both lower performance and higher uncertainty. NLP alone in HLA method also performed moderately (F1 = 0.7479, CI = [0.6890, 0.7985]), while its combination with LLM in SHP (F1 = 0.789, CI = [0.7337, 0.8348]) improved results, reinforcing the benefit of combining augmentation strategies.

A closer examination of robustness reveals that LLM-based augmentation consistently maintains high F1 Scores across both datasets, with relatively narrow confidence intervals. This suggests that the method is less sensitive to variations in dataset characteristics, making it more reliable for practical deployment. In contrast, rule-based and NLP-only methods exhibit wider or more variable confidence intervals, indicating higher susceptibility to fluctuations in data composition. For instance, DSGA augmentation achieves competitive performance on target but shows noticeably lower and more variable F1 Scores on original, highlighting dataset-dependent instability.

Moreover, combinations of LLM with either rule-based or NLP augmentations (SHP and EHA) tend to reduce variability compared to the non-LLM methods alone (DSGA, HLA), demonstrating a stabilizing effect when multiple augmentation strategies are integrated. Nevertheless, it is important to acknowledge that the Wilson confidence intervals provide an approximate measure of uncertainty, and additional statistical validation (e.g., bootstrapping) would be required to fully quantify robustness. Overall, these observations suggest that robustness is closely linked to both the choice of augmentation method and the diversity of the dataset, with LLM-based strategies offering the most consistently reliable performance.

#### 5.2.4 Correlations Between Quality Metrics and Classification Performance

To investigate whether standard quality metrics are predictive of downstream classification gains, Pearson correlation coefficients were computed between BLEU, CHRF, Self-BLEU, Perplexity, and F1 scores achieved on both the original and target evaluation sets. Results show that BLEU and CHRF exhibit strong positive correlations with classification performance, particularly in the original dataset evaluation, where BLEU correlates with F1 at *r* ≈ 0.999, and CHRF at *r* ≈ 0.985. On the target dataset evaluation, correlations remain substantial (BLEU: *r* ≈ 0.802, CHRF: *r* ≈ 0.761), though slightly diminished, likely due to the domain-specific nature of the target set and the effects of second-stage fine-tuning.

Perplexity shows a strong inverse correlation with F1, especially in original dataset (*r* ≈ − 0.6546) and to a moderate degree in target dataset (*r* ≈ − 0.8185), indicating that more fluent and predictable text—as measured by lower perplexity—tends to yield more effective training signals for emotion intensity classification. In contrast, Self-BLEU, a proxy for diversity within the augmented samples, shows weak correlation with performance on MEISD (*r* ≈ 0.308) and moderate correlation on target dataset (*r* ≈ 0.658), suggesting that diversity alone is not a sufficient predictor of downstream effectiveness.

These results imply that task-agnostic metrics such as BLEU, CHRF, and Perplexity, though not designed specifically for emotion-related tasks, can be informative proxies for classification utility in data augmentation scenarios. However, their limitations become apparent when we consider emotional fidelity and stylistic alignment.

#### 5.2.5 Impact of Transfer Learning

The transfer learning setup—pretraining on original and fine-tuning on target dataset—played a crucial role in model generalization. Models trained on original data struggled to transfer effectively (acc = 0.6498), but augmented data improved downstream performance significantly. For example:

- CGA(original) to target: F1 dropped from 0.8816 to 0.8128
- EHA (original) to target: F1 dropped slightly from 0.8352 to 0.8009

While a small drop in performance is expected due to domain shift, the relative stability of CGA and EHA methods demonstrates that augmentation helps bridge the domain gap, improving transferability between emotionally annotated dialogue corpora.

Notably, the models incorporating EDA setups (DSGA, HLA and the SHP) were the only ones that improved after transfer learning from original to target dataset. Specifically, F1 increased from 0.6767 → 0.7479 for HLA and from 0.7361 → 0.7890 for SHP. This suggests that, while these methods did not achieve the best absolute scores on original, they provided greater stability under domain transfer and benefited disproportionately from the target dataset fine-tuning stage.

The hybrid method (SHP), while not consistently superior, achieved a solid middle ground—especially on target dataset (F1 = 0.7890). This suggests that combining diverse augmentation sources may offer marginal robustness, although not outperforming CGA augmentation in this setup.

### 5.3 Error Analysis

To better understand the limitations of our classifier in binary emotion intensity detection, we examined several representative misclassifications. Since the dataset labels indicate only *High* or *Low* emotional intensity, these examples illustrate how different linguistic and contextual cues can mislead the model.

#### 5.3.1 Linguistic Cues Associated with Misclassification

The analysis suggests that certain linguistic patterns systematically mislead the classifier:

- **Conflation of emotional valence and intensity** The model frequently interprets the presence of strong affective words (e.g., *depressed, worried, frustrated*) as indicators of high emotional intensity, even when the overall tone is calm or reflective. For instance, in Case 1 (Table 5.3.2), negative emotion terms co-occurred with a subdued delivery, yet the model predicted “High” with very high confidence.
- **Lack of explicit affect markers in high-intensity cases** Some true “High” intensity examples contained minimal overt emotional vocabulary. Instead, intensity was conveyed through context-dependent signals, such as abrupt sentence endings, ellipses, or implied distress. The model tended to classify these as “Low,” indicating a reliance on explicit lexical cues rather than subtler discourse markers.
- **Narrative detail inflating intensity predictions** In several “Low” labelled cases, long and vivid descriptions of personal experiences appeared to bias the model towards “High” predictions. This suggests that narrative richness and specificity are sometimes mistaken for emotional arousal.

#### 5.3.2 Potential Label Noise

Some misclassifications may be due to inconsistencies in annotation rather than model limitations:

- **Ambiguity in intensity definitions** Case 3 in Table 5.3.2 demonstrates disagreement between the gold label (“Low”) and the emotional intensity suggested by repeated expressions of frustration. Such cases point to differences in annotator interpretation.
- **Context dependency** Short, neutral-sounding utterances (e.g., Case 2) may be labelled “High” when considering the broader conversation but appear “Low” in isolation. Without access to preceding dialogue turns, these instances are inherently difficult for the model to classify correctly.

## 6 Discussion

### 6.1 Domain-Style Alignment and Diminishing Transfer Benefits

The two-stage training strategy—pretraining on a source dataset (original or augmented) followed by fine-tuning on the target dataset—was designed to bridge the gap between domains with distinct communicative and emotional characteristics. Rather than applying simple augmentation, the approach enriched the source data with stylistic and affective cues derived from the target domain, including typical message length, pronoun usage, and emotionally charged vocabulary. This alignment aimed to make the source dataset more compatible with the target domain, improving the model’s adaptability across contexts.

Augmentation itself had a strong impact on performance. As shown in Table 5.4, accuracy on the original dataset increased from 0.6498 to as high as 0.8819 after augmentation, confirming that style-enriched data substantially enhanced representation learning. However, when fine-tuned on the target dataset, performance differences between methods became smaller (Table 5.5). Models based on LLM-generated augmentation (CGA, EHA) achieved the highest source-domain scores but plateaued or slightly declined after fine-tuning, suggesting limited adaptability once transferred.

In contrast, rule-based EDA methods (HLA and SHP) showed the opposite trend: although their initial performance was lower, both improved notably after fine-tuning on the target dataset (HLA +0.0712, SHP +0.0529 F1). This indicates that EDA-based augmentation produced features that were more flexible and responsive to domain adaptation. Among hybrid methods, DSGA performed unexpectedly well, likely due to its balance between stylistic alignment and lexical variation introduced through targeted synonym replacement based on TF–IDF weights.

Overall, these results highlight that data enriched with explicit emotional or stylistic markers can significantly enhance cross-domain transfer. LLM-based augmentation is most effective when in-domain data are scarce, while rule-based or hybrid methods may yield more stable improvements when sufficient target data are available.

### 6.2 Interpretation of Quality Metrics and Their Limitations

The comparative analysis across five augmentation methods highlights that traditional text quality metrics only partially reflect the actual usefulness of synthetic data for downstream emotion intensity classification. While metrics such as BLEU, CHRF, and Perplexity provide valuable insights into surface-level fluency and structural similarity, they do not capture emotional fidelity or domain adaptability, which are critical for affective computing tasks.

#### 6.2.1 Interpreting Transformation Quality

The transformation quality results (Table 5.1) demonstrate that methods leveraging large language models (CGA and EHA) produce synthetically rich and fluent data. The CGA method achieved the highest average quality (0.719±0.161) and near-perfect uniqueness (0.999), indicating strong generative capacity and diversity at the lexical level. However, this apparent superiority in text quality did not fully translate into better classification transfer. Despite their impressive quality metrics, CGA and EHA reached a performance plateau after fine-tuning on the target domain.

In contrast, rule-based approaches incorporating EDA, such as HLA and SHP, initially exhibited lower transformation quality scores (0.328 ± 0.086 and 0.446 ± 0.124, respectively), yet achieved substantial improvements during fine-tuning on the target dataset (HLA +0.0712; SHP +0.0529 F1). This pattern suggests that the more constrained, less “polished” outputs of EDA-based augmentation encouraged models to generalize better across domains. In other words, linguistic imperfection may have introduced beneficial variability that improved adaptability.

The deterministic DSGA and hybrid EHA methods further illustrate this balance. DSGA’s controlled synonym substitution, guided by TF–IDF weighting, produced moderately high-quality yet shorter and more repetitive samples. Nonetheless, its final classification performance was strong, implying that partial stylistic alignment combined with limited variability can stabilize learning. The EHA method, combining 70

#### 6.2.2 Stylistic and Emotional Fidelity

The stylistic consistency results (Table 5.2) offer additional nuance. Interestingly, heuristic methods (HLA, SHP) achieved the highest stylistic alignment with the target domain (average consistency = 0.695), surpassing LLM-based approaches. This outcome reveals that simple lexical manipulations—such as controlled emotional intensity adjustments and pronoun pattern replication—can more precisely reproduce surface-level style than context-heavy LLM outputs. LLM-based models, though fluent, tended to homogenize stylistic cues, which may reduce the distinct emotional tone necessary for empathy-driven text.

#### 6.2.3 Revisiting NLP Metrics

Traditional NLP metrics (Table 5.3) present a complementary yet limited picture. BLEU and CHRF correlated strongly with classification accuracy (Table 5.8), particularly for source-domain evaluation (BLEU *r* = 0.999). However, these correlations weakened after transfer to the target domain (*r* = 0.802), indicating that lexical and structural similarity alone cannot predict success in emotionally nuanced adaptation tasks.

Self-BLEU and lexical diversity metrics (UTR, TTR) exposed another dimension: LLM-based augmentations produced semantically coherent but redundant text (high Self-BLEU, low TTR), while heuristic methods generated more varied, less fluent outputs. Yet, this lexical variety often improved adaptability during fine-tuning, suggesting that internal dataset diversity contributes to generalization even when it slightly reduces BLEU-based “quality.”

Perplexity inversely correlated with F1 (–0.655 and –0.818), confirming that more fluent text generally aids classification. However, beyond a certain threshold of fluency and similarity, additional gains in these metrics yielded diminishing returns. The best-performing EDA-based models demonstrate that moderate linguistic noise may act as a form of regularization, enhancing robustness rather than degrading it.

### 6.3 Limitations of Task-Agnostic Metrics in Emotionally Grounded Tasks

Overall, the findings indicate that conventional quality metrics provide an incomplete representation of augmentation effectiveness in emotion-related tasks. BLEU, CHRF, and PPL capture how well a model reproduces linguistic form, but they overlook what emotional or interpersonal qualities are preserved. The strong yet inconsistent correlations across domains suggest that these metrics primarily reward surface-level similarity rather than functional or affective alignment with communicative goals.

For example, augmentation methods based on EDA (HLA and SHP) achieved higher stylistic consistency with the target dataset than LLM-based augmentations (CGA), even though their BLEU and CHRF scores were lower. This pattern implies that rule-based approaches may better preserve communicative intent and emotional nuance. Conversely, LLM-generated data, while fluent and diverse, tended to homogenize tone and emotional intensity, reducing the richness necessary for empathetic dialogue modeling.

This disconnect underscores a broader limitation: high general-purpose metric scores can mask deficiencies in emotional fidelity or contextual appropriateness, particularly for high-intensity messages where tone modulation is crucial. Hence, while high BLEU or low PPL values signal fluency and grammatical integrity, they cannot reliably capture the subtler aspects of emotional expression or interpersonal tone essential for empathetic response modeling.

Future work should integrate **task-aware evaluators**—such as emotion-style classifiers, empathy scoring models, or expert human annotation—into augmentation pipelines. These complementary evaluators would enable a more comprehensive assessment of both linguistic and affective quality, supporting more reliable and emotionally aligned text generation in affective computing.

### 6.4 Emotional Fidelity Evaluation

To evaluate whether data augmentation and transfer learning effectively preserved emotional meaning, we used emotion intensity classification as a quantitative measure of emotional fidelity. This task assessed how well the augmented samples maintained the intended emotional strength and polarity of the original data after augmentation and domain transfer.

The results demonstrate that augmentation had a substantial positive impact on emotion intensity classification across both datasets. Models trained on augmented data consistently outperformed those trained on the original, non-augmented corpus, confirming that style- and affect-enriched samples supported better affective feature learning. Notably, LLM-based methods such as CGA and EHA yielded the highest overall F1 scores on the source dataset (0.8816 and 0.8352, respectively), indicating strong alignment between linguistic fluency and emotional signal preservation.

However, the comparison between source and target datasets revealed an important nuance. While LLM-generated augmentations achieved the best initial performance, their improvement after fine-tuning was minimal, suggesting that they may have reached a saturation point in emotional representation. In contrast, rule-based EDA methods (HLA and SHP) showed notable gains after transfer learning (+0.0712 and +0.0529 F1), demonstrating higher adaptability and better retention of emotional gradients when exposed to new domain-specific cues.

These findings suggest that the proposed augmentation and transfer learning pipeline not only enhanced linguistic diversity but also successfully preserved and transferred emotional intensity information between domains. Nonetheless, because this evaluation was fully automated, future work should incorporate human or hybrid assessments—for example, expert annotation or emotion-alignment scoring—to better capture subtle nuances of empathy, tone modulation, and communicative intent beyond what model-based classifiers can quantify.

### 6.5 Practical Implications

The findings of this study provide several practical insights for both researchers and practitioners developing affective NLP systems. First, the results confirm that emotion intensity classification can serve as an effective diagnostic tool for assessing whether data augmentation and transfer learning preserve emotional meaning. This offers a scalable alternative to human evaluation, particularly valuable in resource-limited or sensitive domains such as mental health communication.

Second, the experiments show that LLM-based augmentations (such as CGA and EHA) are highly effective when limited in-domain data are available, as they enhance linguistic fluency and emotional fidelity with minimal manual intervention. However, once sufficient target-domain data exist, their relative advantage diminishes, and rule-based or hybrid approaches may provide more stable cross-domain generalization. This finding is particularly relevant for applied settings where domain data gradually accumulate, such as online counseling or emotion-aware chatbots.

Third, the correlation analysis between linguistic quality metrics and downstream F1 performance suggests that task-agnostic measures like BLEU, CHRF, and Perplexity, while imperfect, can still inform model selection during early-stage augmentation. However, they should be complemented with emotion-sensitive metrics or task-specific evaluations to ensure affective and stylistic integrity.

Finally, the methodology of domain-style alignment through targeted augmentation demonstrates a practical pathway for adapting emotion-related models to new conversational domains without exhaustive retraining. By embedding stylistic and affective cues from the target dataset into the augmented source data, the proposed approach effectively “bridges” emotional expression patterns across domains. This enables more robust deployment of affect-aware models in dynamic, real-world contexts where linguistic norms and emotional tone can vary significantly between communities.

### 6.6 Limitations

Although our multi-metric evaluation provides useful signals about augmentation quality, several methodological caveats limit the strength of some claims. First, many of the correlation analyses reported in Section 5 are based on aggregated method-level summaries (*N* ≈ 5 augmentation methods), which can produce unstable Pearson estimates; future work should compute correlations with larger sample counts (e.g., across generation seeds, prompt variants or per-sample measures) and report bootstrap confidence intervals. Second, our binary mapping of intensity levels simplifies the underlying ordinal structure and may obscure subtle improvements; we therefore recommend sensitivity analyses to mapping thresholds and experiments on multi-class/ordinal formulations. Finally, because affective fidelity is central to this task, automatic metrics (BLEU/CHRF/PPL) must be complemented by targeted human evaluations and task-aware emotion-style classifiers to reliably assess whether synthetic samples preserve the intended emotional intensity.

### 6.7 Future Research Directions

While the current study demonstrates the effectiveness of emotion intensity classification as a proxy for evaluating emotional fidelity in augmented data, several research avenues remain open.

1. **Emotion-aware evaluation metrics**. Future work should focus on developing emotion-sensitive automatic metrics that go beyond surface-level similarity. Such metrics could integrate emotion polarity preservation, intensity correlation, or empathy coherence scores derived from specialized classifiers or large-scale human annotation benchmarks. This would enable more nuanced assessment of affective quality in generative augmentation pipelines.
2. **Cross-domain and cross-lingual generalization**. The domain-style alignment strategy presented here can be further tested across languages and communication settings (e.g., peer support, clinical notes, or social media). Exploring multilingual or multicultural adaptation could reveal whether emotional cues transfer consistently across cultural and linguistic boundaries.
3. **Human-in-the-loop augmentation**. Incorporating expert or crowd-sourced feedback during augmentation could refine the affective and stylistic realism of generated data. Hybrid workflows that combine human evaluators with automated scoring may improve both emotional fidelity and data reliability for sensitive domains such as mental health analysis.
4. **Longitudinal validation in downstream systems**. Finally, evaluating the long-term impact of domain-style alignment on deployed systems—such as empathy-enabled chatbots or emotion-aware recommendation engines—would clarify how data augmentation affects real-world emotional engagement and user trust over time.

## 7 Conclusion

This study investigated the effectiveness of various data augmentation strategies for emotion intensity classification in empathetic support dialogues, with a focus on cross-domain adaptation. By evaluating both linguistic quality and emotional fidelity, we demonstrated that augmentation substantially improves classification performance compared to models trained on the original, non-augmented data. LLM-based methods, such as CGA, achieved the highest initial performance and fluency, while rule-based EDA methods (HLA and SHP) proved more adaptable during transfer learning, maintaining or enhancing emotional intensity alignment on target datasets.

The results highlight that standard task-agnostic metrics (BLEU, CHRF, and Perplexity) are informative for assessing linguistic quality but insufficient for capturing affective and stylistic nuances. Emotion intensity classification served as a practical and quantifiable measure of emotional fidelity, validating the success of the augmentation and transfer learning pipeline in preserving affective content.

From a practical perspective, the proposed methodology demonstrates that domain-style alignment through targeted augmentation can bridge source and target datasets, enabling more robust deployment of emotion-aware NLP systems. The findings also indicate that hybrid strategies, which combine LLM-based fluency with rule-based stylistic adjustments, may offer the most balanced approach for both semantic consistency and emotional expressiveness.

Future work should focus on developing emotion-sensitive evaluation metrics, exploring the evolution of emotion intensity across full multi-turn conversations, and extending the methodology to cross-domain and multilingual settings. Additionally, incorporating human-in-the-loop validation could further ensure nuanced affective quality, supporting safe and effective deployment in real-world empathetic dialogue applications.

Overall, this study provides a comprehensive framework for evaluating and improving affective data augmentation, offering actionable insights for researchers and practitioners aiming to build emotionally-aware NLP systems capable of generalizing across domains.

## Data Availability

All data produced in the present work are contained in the manuscript

## Notes

### Competing Interest Statement

The authors have declared no competing interest.

### Funding Statement

This study did not receive any funding

